# Adiposity distribution and risks of twelve obesity-related cancers: a Mendelian randomization analysis

**DOI:** 10.1101/2025.01.10.25320324

**Authors:** Emma Hazelwood, Lucy J. Goudswaard, Matthew A. Lee, Marina Vabistsevits, Dimitri J. Pournaras, Hermann Brenner, Daniel D Buchanan, Stephen B Gruber, Andrea Gsur, Li Li, Ludmila Vodickova, Robert C. Grant, N. Jewel Samadder, Nicholas J. Timpson, Marc J. Gunter, Benjamin Schuster-Böckler, James Yarmolinsky, Tom G. Richardson, Heinz Freisling, Neil Murphy, Emma E. Vincent

**Affiliations:** MRC Integrative Epidemiology Unit, University of Bristol, Bristol, UK; Population Health Sciences, Bristol Medical School, University of Bristol, Bristol, UK; Nutrition and Metabolism Branch, International Agency for Research on Cancer (IARC-WHO), Lyon, France; Clinical and Biomedical Sciences, University of Exeter, Exeter, UK; Department of Upper GI and Bariatric/Metabolic Surgery, North Bristol NHS Trust, Southmead Hospital, Bristol, UK; Division of Clinical Epidemiology and Aging Research, German Cancer Research Center (DKFZ), Heidelberg, Germany; Division of Preventive Oncology, German Cancer Research Center (DKFZ) and National Center for Tumor Diseases (NCT), Heidelberg, Germany; German Cancer Consortium (DKTK), German Cancer Research Center (DKFZ), Heidelberg, Germany; Colorectal Oncogenomics Group, Department of Clinical Pathology, Melbourne Medical School, The University of Melbourne, Parkville, Australia; University of Melbourne Centre for Cancer Research, The University of Melbourne, Parkville, Australia; Genomic Medicine and Family Cancer Clinic, The Royal Melbourne Hospital, Parkville, Victoria, Australia; Department of Medical Oncology & Therapeutics Research and Center for Precision Medicine, City of Hope National Medical Center, Duarte, California, USA; Center for Cancer Research, Medical University of Vienna, Vienna, Austria; Department of Family Medicine, University of Virginia, Charlottesville, Virginia, USA; Institute of Biology and Medical Genetics, First Faculty of Medicine, Charles University, Prague, Czech Republic; Faculty of Medicine and Biomedical Center in Pilsen, Charles University, Pilsen, Czech Republic; Division of Medical Oncology and Hematology, Princess Margaret Cancer Centre, Toronto, Ontario, Canada; Division of Gastroenterology, Mayo Clinic, Phoenix, Arizona, USA; Department of Epidemiology and Biostatistics, School of Public Health, Imperial College London, UK; Ludwig Institute for Cancer Research, University of Oxford, Oxford, UK; Translational Health Sciences, Bristol Medical School, University of Bristol, Bristol, UK

## Abstract

**Background:** There is convincing evidence that overall adiposity (as measured by body mass index) increases the risks of several cancers. Whether there are similar relationships between these cancers and the distribution of adiposity is unclear.

**Methods:** In the absence of well-powered individual studies we utilised two-sample Mendelian randomization (MR) to examine causal relationships of five adiposity distribution traits (abdominal subcutaneous adipose tissue; ASAT, visceral adipose tissue; VAT and gluteofemoral adipose tissue; GFAT, liver fat, and pancreas fat) with the risks of 12 obesity-related cancers (endometrial, ovarian, breast, colorectal, pancreas, multiple myeloma, liver, kidney (renal cell), thyroid, gallbladder, oesophageal adenocarcinoma, and meningioma) and cancer subtypes/subsites. We then used multivariable MR to investigate whether plausible molecular intermediates could potentially be mediating the relationships identified. We used the largest available GWAS from European populations for all traits (sample size across all GWAS ranged from 8,407 to 728,896 (median: 57,249); cancer GWAS ranged from 279 to 133,384 cases (median: 4,532) and 3,456 to 727,247 controls (median: 68,802)).

**Results:** We found evidence that higher genetically predicted ASAT increased risks of endometrial cancer (inverse variance-weighted (IVW) odds ratio (IVW OR) per standard deviation (SD) higher genetically predicted ASAT = 1.79, 95% confidence interval (CI) = 1.18 to 2.71), liver cancer (IVW OR per SD higher genetically predicted ASAT = 3.83, 95% CI = 1.39 to 10.53) and oesophageal adenocarcinoma (IVW OR per SD higher genetically predicted ASAT = 2.34, 95% CI = 1.15 to 4.78). Conversely, we found evidence that higher genetically predicted GFAT decreased risks of breast cancer (IVW OR per SD higher genetically predicted GFAT= 0.77, 95% CI = 0.62 to 0.97) and meningioma (IVW OR per SD higher genetically predicted GFAT = 0.53, 95% CI = 0.32 to 0.90). We also found evidence for an effect of higher genetically predicted VAT and liver fat on increased liver cancer risk (IVW ORs per SD higher genetically predicted adiposity trait = 4.29 and 4.09, 95% CIs = 1.41 to 13.07 and 2.29 to 7.28, respectively). Multivariable MR analyses suggested that traits related to insulin signalling, sex hormones, and inflammation may play important roles in mediating the effects of adiposity distribution on obesity-related cancers.

**Conclusions:** Our analyses provide novel insights into the variability of the effect of adiposity distribution on cancer risk, with respect to both adiposity trait and cancer type, which would not be possible in a conventional observational analysis given the lack of available samples with all required traits measured. These findings demonstrate that adipose tissue at different anatomical locations may have differential effects on adiposity-related molecular traits. These insights enhance our understanding of the complex relationship between adiposity and cancer risk and highlight the importance of adipose tissue distribution alongside maintaining a healthy weight overall for cancer prevention.

## Introduction

The prevalence of obesity is increasing worldwide, having doubled in more than 70 countries over the past four decades with 4 billion individuals estimated to be living with obesity by 2035[1,2]. Obesity is known to increase the risk of cancer, which is the second most common cause of death worldwide[3]. In 2016, an International Agency for Research on Cancer (IARC) report concluded that there is sufficient evidence to support an association of obesity with risks of at least 13 cancers - endometrial, ovarian, breast, colorectal, pancreas, multiple myeloma, liver, kidney (renal cell), thyroid, gallbladder, oesophageal adenocarcinoma, gastric cardia and meningioma[4]. A growing body of evidence shows that effective obesity care can lead to a reduction in cancer risk[5–12]. However, the importance of the anatomical distribution of adiposity in the obesity-cancer risk relationship is not fully understood.

Adipose tissue, primarily consisting of adipocytes, is distributed throughout the body and plays a crucial role in energy storage and metabolism[13,14]. Additionally, it functions as an endocrine organ, secreting hormones and circulatory factors[15]. Each deposit originates from a distinct fat progenitor and has a unique microenvironment and cytological make up[16,17]. For instance, visceral adipose tissue has an important role in the secretion of inflammation-related adipokines and growth factors, whereas subcutaneous adipose tissue is the predominant source of circulating oestrogen in men and post-menopausal women[18,19]. Furthermore, each deposit interacts with different organs and biological structures proximal to the adipose tissue[20].

Body mass index (BMI) is often utilised as a proximal measure of adiposity in epidemiological studies. As a simple anthropometric derivative (height for stature) correlated with many other body composition traits, BMI is useful in large-scale population studies. However, BMI cannot capture the distribution of adipose tissue throughout the body [21–23]. Recent research has revealed that more specific measures, including abdominal subcutaneous, visceral, and gluteofemoral adipose tissue (ASAT, VAT, and GFAT, respectively), as well as the fat content of certain organs, in particular the liver and pancreas, have been shown to have differential roles in the effect of adiposity on risk of cardiometabolic outcomes[21,24,25]. Such findings have led to important updates in clinical guidelines, for example, the European Association for the Study of Obesity (EASO) framework which now stresses that “BMI alone is insufficient as a diagnostic criterion, and body fat distribution has a substantial effect on health.”[26]. However, in contrast to cardiometabolic outcomes, the adiposity distribution-cancer relationship has thus far only been evaluated in observational settings where results may reflect residual confounding and/or reverse causation, meaning the causality of these relationships is unknown. In addition, the effects of adiposity distribution on circulating molecular traits – such as sex hormones, inflammatory biomarkers, and metabolic features, including those related to insulin signalling and fatty acid metabolism – in relation to cancer risk are also understudied. Therefore, the mediators of the potential causal relationships between adiposity distribution traits and cancer risks remain unclear. Given the rise in obesity worldwide and its effect on risks of multiple cancer types, investigating the roles of adiposity distribution and related mediating traits in these relationships thus represents a clinically important step forward in deepening our understanding of which individuals are most at risk for different cancer types, and which interventions are likely to be most effective for cancer prevention.

Currently, there is a lack of large-scale data in which adiposity distribution, cancer-related molecular traits, and cancer status are all measured in the same individuals. In order to evaluate evidence for causal effects of different adiposity distribution traits with the risks of obesity-related cancers, we employed two-sample Mendelian randomization (MR), a genetic epidemiological approach which uses genetic instruments as proxies to investigate causal relationships between traits[27,28]. We used MR to (i) evaluate the effects of five adiposity traits on risks of 12 obesity-related cancers; (ii) evaluate the effects of the same adiposity traits on cancer-related molecular traits; (iii) evaluate the effects of adiposity distribution-related molecular traits on cancer risk; and (iv) estimate the proportion of the effects of adiposity distribution traits on cancer risks that may be mediated by the molecular traits identified.

## Methods

We performed four analyses in sequence (**Figure 1**). First, we performed MR to evaluate the causal effect of five adiposity distribution traits and BMI on the risks of 12 obesity-related cancers. Second, we performed an MR analysis of the effect of adiposity traits on circulating levels of various molecular traits which are potential mediators of the effects of adiposity measures on cancer risk. Third, we performed an MR analysis of molecular traits (those for which there was evidence of a causal effect of adiposity on) to risks of the 12 obesity-related cancers. Analyses two and three represent a two-step MR approach to identify potential mediating traits in these relationships[29,30]. Finally, we performed multivariable MR to evaluate and quantify the potential mediating role of these traits in the causal effects of adiposity distribution on risks of obesity-related cancers.

**Figure 1.**
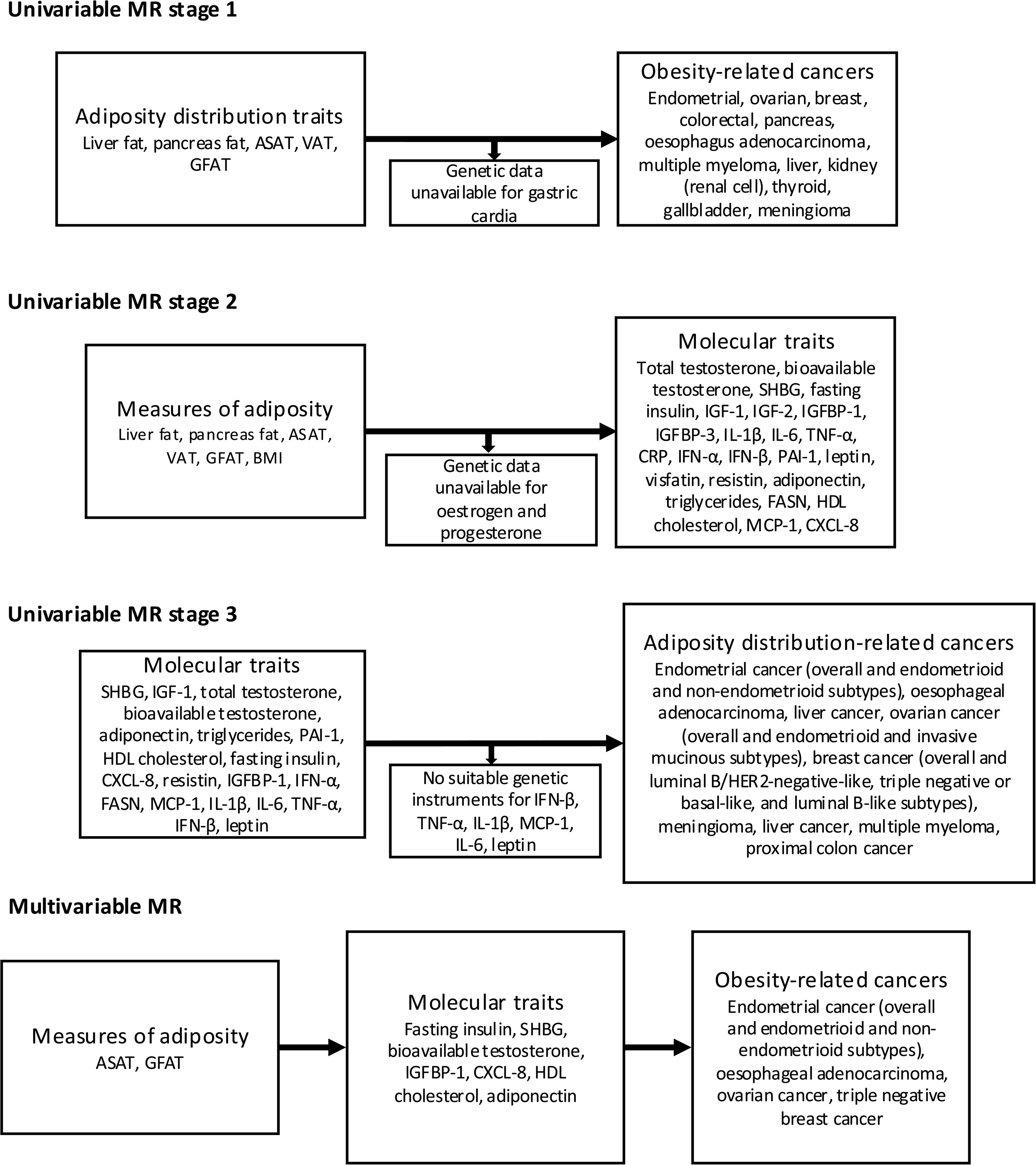
Flowchart detailing analysis plan.

### Study populations

**Table 1** shows the source of genetic data for each trait used in this study. In some instances, there was population overlap between GWAS, in these cases analyses were repeated using GWAS from independent populations for the adiposity, molecular, and cancer traits. For six cancers (**Supplementary table 1**), data from UK Biobank were meta-analysed with data from FinnGen (discussed below). Given the adiposity data were also generated using UK Biobank, it is likely that high levels of sample overlap are present for MR analyses using these data. As such, particularly in the presence of weak instruments, estimates may be biased[31,32]. However, although weak instrument bias is a linear function of the degree of overlap, incorporating a larger sample size (and often thereby increasing the degree of sample overlap) can also increase the strength of the instruments, meaning there is a trade-off between instrument strength and sample overlap in two-sample MR analyses.

Summary genetic data for all five measures of adiposity distribution (unadjusted for BMI in order to avoid collider bias[33,34]) were obtained from genome-wide association studies (GWAS) using UK Biobank[21,24]. UK Biobank enrolled over 500,000 individuals between the ages of 40 and 69 years between 2006 and 2010[35]. Summary genetic data for the adiposity distribution traits were obtained from GWAS of 38,965 UK Biobank participants who underwent magnetic resonance imaging (MRI) scans, followed by quantification of the adiposity measures using a deep learning model trained on body MRI images[21,24]. Prior to performing MR analyses, pairwise genetic correlation of the adiposity distribution traits and BMI was calculated using LD score regression[36,37] (RG = 0.2 to 0.8, median = 0.5, **Supplementary figure 1**).

Overall, 12 of the 13 cancers identified in the IARC report had summary genetic data available[4] (**Table 1)**; data were unavailable for gastric cardia (also known as upper stomach) cancer. This cancer often overlaps with oesophageal adenocarcinoma (for which data were available), as many cases are junctional involving the cardia. Where available, summary genetic association data were obtained from cancer GWAS consortia or large-scale meta-analyses. Where such data were not available, a meta-analysis was conducted using summary genetic data from UK Biobank and FinnGen (**Table 1)**. Subtype- or anatomical subsite-specific data were included where available. See **Supplementary methods** for more information on the cancer GWAS included in this analysis.

### Meta-analysis of UK Biobank and FinnGen cancer cases

For kidney (renal-cell) cancer, thyroid cancer, multiple myeloma, liver cancer, and gallbladder cancer, meta-analyses were performed using METAL v2011-03-25 software. We used the software to combine test statistics and standard errors and controlled for population stratification (using the METAL genomic control feature) twice – once for each GWAS individually, and again for the meta-analysed results, as recommended in the METAL documentation (for more information see: https://genome.sph.umich.edu/wiki/METAL_Documentation)[38]. We then filtered the resulting meta-analysis results to remove any single-nucleotide polymorphisms (SNPs) with a heterogeneity *P*-value < 0.05. This may be particularly pertinent for meta-analyses involving FinnGen given that many of the participants in this cohort were hospital-based recruits, which can induce selection bias in case-control outcomes.

### Identification of potentially mediating molecular traits

We evaluated whether the adiposity measures had distinct molecular effects, and whether these molecular traits were in turn mediating the effects of the adiposity measures on cancer risk. To this end, we identified 26 molecular traits which plausibly mediate the effect of adiposity on cancer risk. This *a priori* list of traits was identified from the 2018 World Cancer Research Fund (WCRF) Continuous Update Project (CUP) report titled “Body fatness and weight gain and the risk of cancer”[39]. Specifically, we included all molecular traits listed in “Appendix 2: Mechanisms” of this report (see **Supplementary methods** for more information). Suitable summary genetic data were available for 24 of these 26 traits (all except oestrogen and progesterone). Additional information on the GWAS used for these traits, including sample sizes and ancestry, is available in **Table 1**.

### Mendelian randomization analyses

We performed MR to evaluate evidence for a causal effect of the exposures on the outcomes. MR can result in unbiased estimates of causal effects if the following assumptions are met: (i) the instrument strongly associates with the exposure; (ii) there is no confounding of the instrument-outcome relationship; and (iii) the instrument only affects the outcome through the exposure[40]. Two-sample MR has the additional assumption that the samples used to obtain SNP-exposure and SNP-outcome associations must be representative of the same underlying population[41]. The STROBE-MR reporting guidelines have been followed throughout this manuscript (**Supplementary note)**[42,43].

To construct genetic instruments for MR analyses, we obtained SNPs strongly (*P* < 5 × 10^−8^) and independently (*r*^2^ < 0.001) associated with each trait. For protein traits (i.e. IGFBP-1, adiponectin, IFN-β, IL-6, PAI-1, resistin, TNF-α, IL-1β, visfatin, IGF-1, C-reactive protein, MCP-1, leptin, and FASN), we restricted genetic instruments to all remaining *cis*-acting SNPs (those in or within a 1MB window around the gene coding region) after clumping to minimise possible horizontal pleiotropy. Using this approach, no suitable genetic instruments were available for IFN-β, TNF-α, IL-1β, MCP-1, leptin, or IL-6, meaning these traits were not instrumented as exposures in MR analyses. It should be noted that previous analyses have employed more comprehensive approaches to identify genetic instruments to proxy some of these traits, for instance by using alternative GWAS in the case of leptin or using SNPs in or near a relevant receptor (and weighting by their effect on C-reactive protein) in the case of IL-6[44,45]. Given the large number of traits included in these analyses, we maintained a systematic approach to instrument construction as described above and did not implement previously described approaches to identify instruments for FN-β, TNF-α, IL-1β, MCP-1, leptin, and IL-6. For sex hormone and related traits (i.e. total testosterone, bioavailable testosterone, and sex hormone-binding globulin (SHBG)), genetic instruments were constructed using sex-specific GWAS. The genetic instruments used for all traits are shown in **Supplementary table 2**.

Where there was a single genetic instrument available for a trait, the Wald ratio and delta method were used to calculate effect estimates and approximate standard errors, respectively[46]. Where there were multiple genetic instruments available, inverse-variance weighted (IVW) multiplicative random-effects models were applied. Where there were more than 10 genetic instruments available for a trait, weighted median estimation and weighted mode estimation were used to evaluate evidence for horizontal pleiotropy (a violation of assumption (iii) of MR)[47–50]. As a heuristic, any analyses with a *P*-value below 0.05 was classed as having evidence for an effect. In addition, we categorised any analysis which passed a Bonferroni correction accounting for multiple testing (*P* < 0.05/12 cancer types; 0.05/24 molecular traits; 0.05/21 unique adiposity-cancer pairs; or 0.05/10 multivariable MR analyses) as having “strong” evidence for an effect.

F-statistics were calculated (as R^2^ x (N-1)/((1-R^2^) x k), where k is the number of genetic instruments and N is sample size of the exposure GWAS) for all SNPs for each individual exposure; any SNP with an F-statistic < 10 was excluded (and a mean was calculated for each exposure) prior to conducting MR analyses, to assess potential for weak instrument bias (a violation of assumption (i) of MR)[51]. In addition, Steiger filtering was performed prior to MR analyses, with any genetic instruments explaining more variance in the outcome than the exposure excluded[52] (see **Supplementary methods** for more information). Post-hoc power calculations were performed for all MR analyses evaluating evidence for a causal effect of adiposity measures on cancer risk with a series of odds ratios (IVW ORs) to produce power curves demonstrating estimated power in our analyses for a given magnitude of causal effect of the adiposity measure on cancer risk. It should be noted that these magnitudes of causal effect are subject to the normalisation procedure conducted in the measures of adiposity GWAS meaning it is unclear what a biologically relevant magnitude would be (**Supplementary figure 2**).

We employed several sensitivity analyses to evaluate the reliability of our results. We repeated all analyses avoiding sample overlap by using summary-level data for the exposure or outcome that did not contain UK Biobank, as outlined in **Supplementary methods**. Additionally, for any MR analyses where at least one of the traits is likely to have sex-specific genetic architecture (e.g. sex hormones and related traits), or where one GWAS was limited to a specific sex (e.g. breast and endometrial cancer), we repeated analyses with sex-specific GWAS where available for both exposure and outcome, regardless of the strength of evidence in the sex-combined analyses. **Table 1** shows further details on the GWAS used for these sensitivity analyses. Note that sex-specific data were used for all sex hormone traits throughout, as described above.

As an additional sensitivity analysis, we aimed to perform multivariable MR to formally evaluate evidence for distinct roles of different adipose depots, particularly those which had similar effect estimates for cancer risk in the univariable MR analyses. However, combining all five adipose depots in a single multivariable MR model resulted in conditionally weak instruments (all conditional F-statistics <10, median = 5; see **Supplementary table 3**). Therefore, we also calculated conditional F-statistics for all pairwise combinations of the traits. We then performed multivariable MR for any pairs of adiposity traits where both had conditional F-statistics > 10, and where both traits had at least nominal evidence (*P* < 0.05) for an effect on the same cancer in univariable analyses.

### Mediation analysis

For all potential mediating molecular traits, we performed multivariable MR[53–55] to evaluate evidence for a mediating role and to estimate the proportion of the total effect of the adiposity measure on cancer risk being mediated. This was calculated using the product of coefficients method, after generating estimates of the direct effect (i.e. the remaining effect of the exposure on the outcome when the effect of the candidate mediator on the outcome has been adjusted for) and indirect effect (i.e. the effect of the exposure on the outcome through the candidate mediator)[56]. Standard errors for the proportion mediated were calculated using the delta method[57]. Conditional F-statistics using an assumed covariance of zero were calculated to evaluate the potential presence of weak instrument bias in these analyses[54,58].

## Results

### Evaluating the effect of measures of adiposity on risk of obesity-related cancers

All results are given per standard deviation (SD) increase in the exposure with 95% confidence intervals (CIs); where continuous traits were inverse rank normally transformed prior to genome-wide analysis we assume the distribution of the traits prior to transformation was normal and therefore we consider these to be normalised SD units. There was evidence (*P*-value < 4.17 x 10^-3^; 0.05/12 cancer types) for an effect of higher genetically predicted liver fat on liver cancer risk (IVW OR = 4.09, CI = 2.29 to 7.28), higher genetically predicted pancreas fat on endometrioid ovarian cancer risk (IVW OR = 1.99, CI = 1.37 to 2.90), and higher genetically predicted ASAT on luminal B/HER2-negative-like (IVW OR = 0.54, CI = 0.40 to 0.73) and triple negative or basal-like breast cancer risk (IVW OR = 0.43, CI = 0.26 to 0.71; **Figure 2**; **Table 2)**. Without multiple testing correction, there was also evidence for a causal effect of higher genetically predicted ASAT, VAT, GFAT, and pancreas fat on risk of several further cancer types (**Figure 2**; **Table 2)**. The results of the MR analysis evaluating the causal effect of BMI on risk of the 12 cancers and subtypes are provided in **Supplementary table 4**.

**Figure 2.**
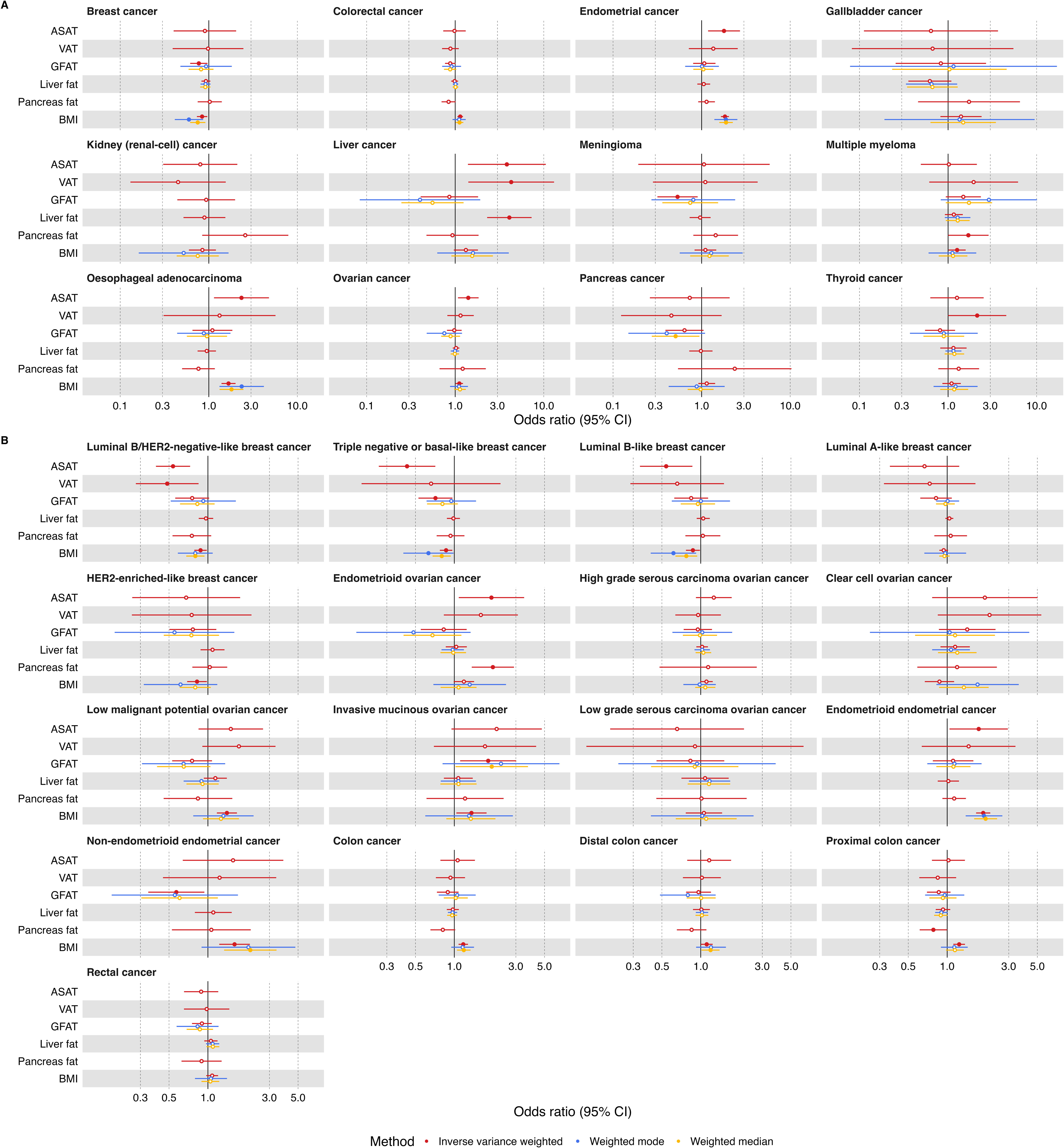
Univariable MR results examining the effect of measures of adiposity on risk of obesity-related cancers, (A) overall and (B) by subtypes. Odds ratios shown are given as one SD increase in adiposity measure. Open/closed circles indicate the *P*-value did not/did meet the evidence threshold (*P*-value < 0.05) respectively. ASAT = adipose subcutaneous adipose tissue; VAT = visceral adipose tissue; GFAT = gluteofemoral adipose tissue; BMI = body mass index.

Repeating initial MR analyses of VAT on risk of luminal B/HER2-negative-like breast cancer using female-specific data resulted in a different direction of effect to those derived from the primary (i.e. sex-combined VAT) analysis. In all other sensitivity analyses, estimates derived from those examining the effect of sample overlap and sex-specific data were in a consistent direction with those derived from the primary analyses, although some 95% CIs crossed the null (**Supplementary figures 3 and 4** , **Supplementary tables 5 and 6**).

Based on these results, we prioritised adiposity trait and cancer pairs to take forward for subsequent analyses. ASAT was paired with: luminal B/HER2-negative-like breast cancer, triple negative or basal-like breast cancer, endometrial cancer, liver cancer, luminal B-like breast cancer, ovarian cancer, oesophageal adenocarcinoma, endometrioid ovarian cancer, and endometrioid endometrial cancer. GFAT was paired with: invasive mucinous ovarian cancer, meningioma, breast cancer, non-endometrioid endometrial cancer, triple negative or basal-like breast cancer, and invasive mucinous ovarian cancer. VAT was paired with: liver cancer and thyroid cancer. Liver fat was paired with liver cancer. Pancreas fat was paired with: endometrioid ovarian cancer, multiple myeloma, and proximal colon cancer.

In the sensitivity analysis using multivariable MR to evaluate evidence for distinct roles of different adipose depots, the pairwise combinations where both adiposity traits had conditional F-statistics > 10 were: ASAT and liver fat; GFAT and liver fat; GFAT and pancreas fat; and liver fat and pancreas fat (**Supplementary table 7**). Of these adiposity traits, ASAT and liver fat only had evidence for an effect on the same cancer in the univariable MR analyses, in that there was evidence for an effect of both traits on the risk of liver cancer (**Figure 2)**. Thus, we performed a multivariable MR to establish whether the ASAT and liver fat genetic instruments were capturing shared or distinct biological mechanisms in the association with risk of liver cancer. In the multivariable MR analysis examining their role on liver cancer risk, the effect estimates for both ASAT and liver fat were attenuated towards the null (**Supplementary** Figure 7**; Supplementary table 8**).

### Evaluating the effect of measures of adiposity on molecular traits

All results are given as an SD change in the outcome per SD increase in the exposure with 95% CIs unless units are otherwise stated in **Table 1**. There was strong evidence for a causal effect of ASAT on leptin (IVW beta = 0.71, CI = 0.53 to 0.89), sex hormone-binding globulin (SHBG; IVW beta = -0.27, CI = -0.38 to -0.15,), IGFBP-1 (IVW beta = -0.40, CI = -0.63 to -0.17), IGF-1 (IVW beta = -0.14, CI = -0.22 to -0.06), total testosterone (IVW beta = -0.07, CI = -0.11 to -0.03); VAT on leptin (IVW beta = 1.10, 95% CI = 0.81 to 1.38); GFAT on triglycerides (IVW beta = -0.40, 95% CI = -0.56 to -0.24), IGF-1 (IVW beta = -0.22, 95% CI = -0.34 to -0.10), HDL cholesterol (IVW beta = 0.31, 95% CI = 0.12 to 0.50); and liver fat on adiponectin (IVW beta = -0.40, 95% CI = -0.62 to -0.19) (**Figure 3**; **Table 3**). There was evidence of potential horizontal pleiotropy for the association between GFAT and triglycerides in that the effect estimate derived using a weighted mode model was in the opposite direction of effect from those derived using the inverse variance weighted and weighted median models. There was also evidence for a causal effect of higher genetically predicted ASAT, GFAT, pancreas fat and liver fat on a 18 further molecular traits (**Figure 3**; **Table 3)**.

**Figure 3.**
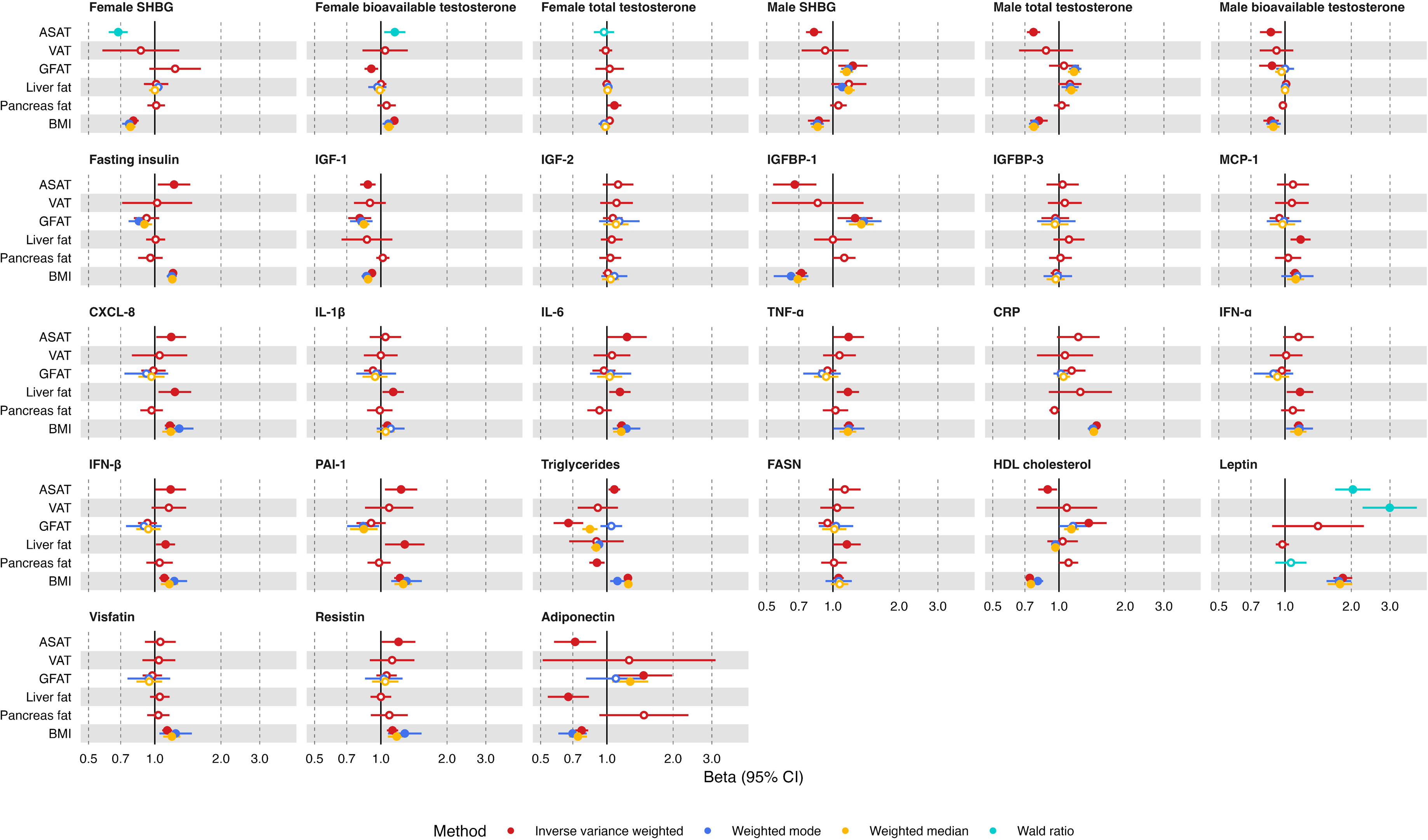
Univariable MR results examining the effect of measures of adiposity on potential molecular mediators of the effect of adiposity on cancer risk. Betas shown are given as one SD increase in adiposity measure and inverse-normal transformed nmol/L total testosterone; natural log transformed nmol/L bioavailable testosterone; inverse rank normal transformed SD SHBG; natural log transformed pmol/L fasting insulin; nmol/L IGF-1; SD IGF-2; SD IGFBP-1; SD IGFBP-3; SD MCP-1; SD CXCL-8; SD IL-1B; SD IL-6; SD TNF-a; mg/L CRP; SD IFN-a; SD IFN-B; SD PAI-1; SD triglycerides; SD FASN; SD (0.38 mmol/L) HDL cholesterol; SD leptin; SD visfatin; SD resistin; SD adiponectin. Open/closed circles indicate the *P*-value did not/did meet the evidence threshold (*P*-value < 0.05) respectively. ASAT = adipose subcutaneous adipose tissue; VAT = visceral adipose tissue; GFAT = gluteofemoral adipose tissue; SHBG = sex hormone-binding globulin; IGF = insulin-like growth factor; IGFBP = IGF binding protein; MCP = monocyte chemotactic protein; CXCL = C-X-C motif chemokine ligand; IL = interleukin; TNF = tumour necrosis factor; CRP = C-reactive protein; IFN = interferon; PAI = plasminogen activator inhibitor; FASN = fatty acid synthase; HDL = high-density lipoprotein.

In sensitivity analyses examining the effect of sample overlap and sex-specific data, estimates were in a consistent direction with those derived from the primary analysis although some 95% CIs crossed the null (**Supplementary figures 3 and 5** , **Supplementary tables 5 and 6**). The single genetic instrument for female-specific VAT was not available (and no suitable proxies could be identified) for the MR analyses of VAT on female-specific sex hormones, so for these analyses the genetic instrument from the sex-combined VAT GWAS was used.

Based on these results, we prioritised adiposity trait and molecular trait pairs to take forward for subsequent analyses. ASAT was paired with: SHBG (female and male), IGFBP-1, IGF-1, total testosterone (male), bioavailable testosterone (female and male), adiponectin, triglycerides, PAI-1, HDL cholesterol, fasting insulin, CXCL-8, and resistin.

GFAT was paired with IGF-1, HDL cholesterol, adiponectin, IGFBP-1, SHBG (male), and bioavailable testosterone (female and male). Liver fat was paired with adiponectin, CXCL-8, PAI-1, IFN-α, and FASN. Pancreas fat was paired with triglycerides and total testosterone (female).

### Evaluating the effect of molecular traits on risk of obesity-related cancers

All cancers and molecular traits with evidence of association with an adiposity measure were investigated for associations with one another. All results are given per SD increase in the exposure with 95% CIs unless units are otherwise stated in **Table 1**. In these analyses, there was strong evidence (i.e. passing Bonferroni correction) for a causal effect of SHBG on endometrial cancer risk (IVW OR = 0.83, 95% CI = 0.76 to 0.90) and endometrioid endometrial cancer risk (IVW OR = 0.82, 95% CI = 0.74 to 0.89), fasting insulin on endometrial cancer risk (IVW OR = 2.75, 95% CI = 1.58 to 4.78) and endometrioid endometrial cancer risk (IVW OR = 3.08, 95% CI = 1.57 to 6.01), and HDL cholesterol on triple negative or basal-like breast cancer risk (IVW OR = 1.16, 95% CI = 1.06 to 1.26).

There was also evidence for a causal effect of total testosterone on risk of endometrial cancer; IGF-1 and leptin on risk of luminal B/HER2-negative-like breast cancer; adiponectin on risk of triple negative or basal-like breast cancer; total testosterone on risk of luminal B-like breast cancer; total testosterone on risk of endometrioid endometrial cancer; and SHBG on risk of non-endometrioid endometrial cancer (**Figure 4**; **Table 4**). In general, effect estimates of consistent direction were found in the weighted median and mode models. The genetic instruments for IGF-1 were not available in the GWAS for ovarian cancer, invasive mucinous ovarian cancer, and endometrioid ovarian cancer and no suitable proxy SNPs were available, meaning the causal effect if IGF-1 on these cancers could not be estimated.

**Figure 4.**
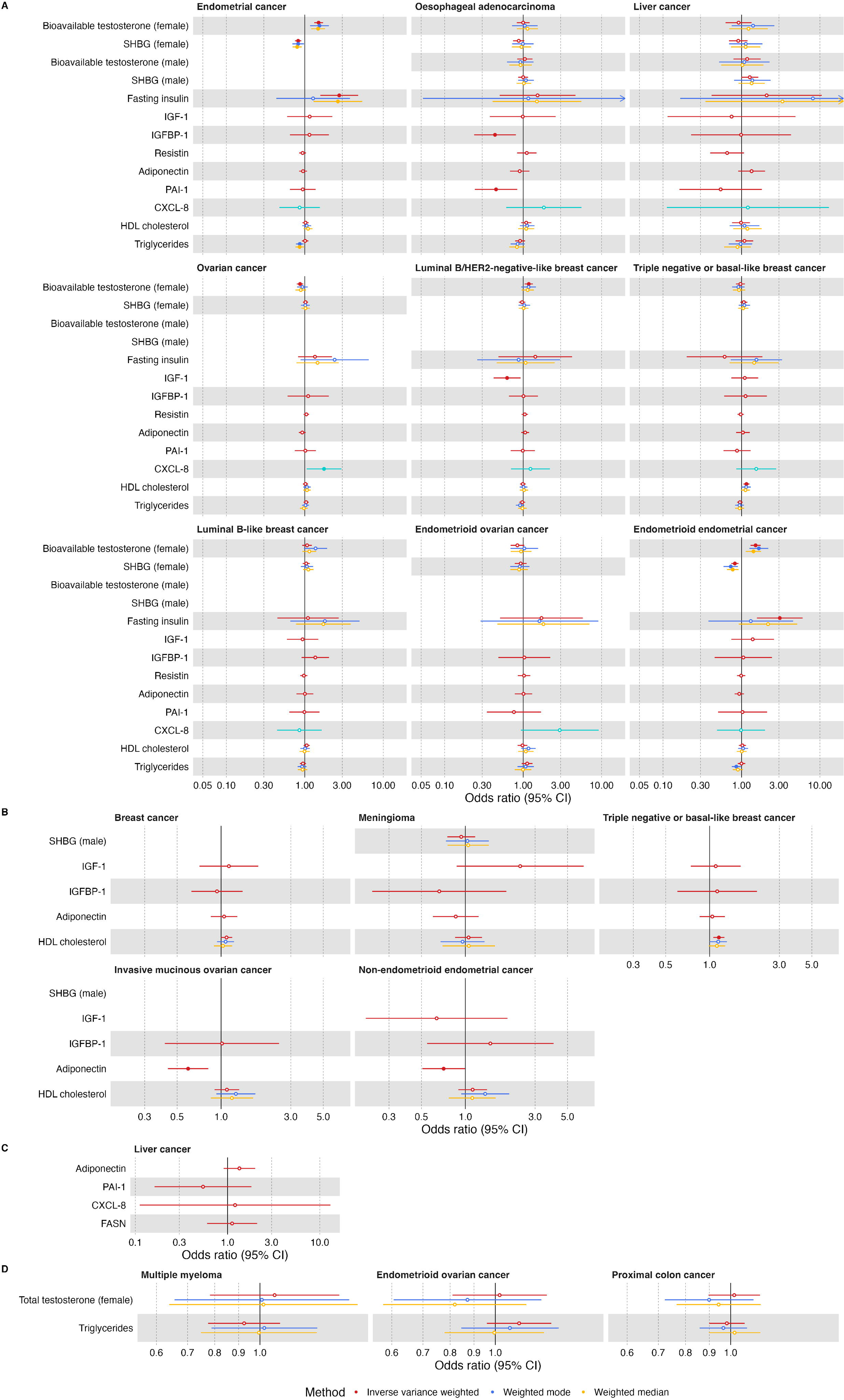
Univariable MR results examining the effect of potential molecular mediators of the effect of adiposity distribution on cancer risk. (A) Molecular traits and cancers which were found to be affected by ASAT in earlier MR analyses; (B) Molecular traits and cancers which were found to be affected by GFAT in earlier MR analyses; (C) Molecular traits and cancers which were found to be affected by liver fat in earlier MR analyses; (D) Molecular traits and cancers which were found to be affected by Pancreas fat in earlier MR analyses. Odds ratios shown are given as increase of one inverse-normal transformed nmol/L total testosterone; inverse rank normal transformed SD SHBG; natural log transformed pmol/L fasting insulin; nmol/L IGF-1; SD IGFBP-1; SD resistin; SD adiponectin; SD PAI-1; SD CXCL-8; SD (0.38 mmol/L) HDL cholesterol; SD triglycerides; natural log transformed nmol/L bioavailable testosterone. Open/closed circles indicate the *P*-value did not/did meet the evidence threshold (*P*-value < 0.05) respectively. ASAT = adipose subcutaneous adipose tissue; VAT = visceral adipose tissue; GFAT = gluteofemoral adipose tissue; SHBG = sex hormone-binding globulin; IGF = insulin-like growth factor; IGFBP = IGF binding protein; PAI = plasminogen activator inhibitor; CXCL = C-X-C motif chemokine ligand; HDL = high-density lipoprotein.

In sex-specific sensitivity analyses, estimates were in a consistent direction with those derived from the primary analysis although some 95% CIs crossed the null (**Supplementary table 6, Supplementary figure 5**). There was not any sample overlap for the MR analyses of molecular traits on cancer risks where evidence for an effect was observed. For the following MR analyses, the direction of the effect of the molecular trait on the cancer risk was not consistent with the direction of the effect of the adiposity distribution trait on the molecular trait and the cancer risk (e.g. if the adiposity trait increased levels of the molecular trait and increased risk of a cancer, the molecular trait would need to increase risk of the cancer): bioavailable testosterone (female) on risks of ovarian and luminal B/HER2-negative-like breast cancer, PAI-1 on risk of oesophageal adenocarcinoma, IGF-1 on risk of luminal B/HER2-negative-like breast cancer, HDL cholesterol on risk of triple negative breast cancer, and adiponectin on risk of invasive mucinous ovarian cancer.

Given these results, the following adiposity trait-molecular trait-cancer trios were taken forward for mediation analyses: ASAT-fasting insulin-endometrial cancer; ASAT-fasting insulin-endometrioid endometrial cancer; ASAT-SHBG (female)-endometrial cancer; ASAT-SHBG (female)-endometrioid endometrial cancer; ASAT-bioavailable testosterone (female)-endometrial cancer; ASAT-bioavailable testosterone (female)-endometrioid endometrial cancer; ASAT-IGFBP-1-oesophageal adenocarcinoma; ASAT-CXCL-8-ovarian cancer; ASAT-HDL cholesterol-triple negative breast cancer; and GFAT-adiponectin-non-endometrioid endometrial cancer.

### Multivariable MR mediation analysis

For all potential biological mechanisms identified (i.e. where we found evidence for an effect of an adiposity trait on a molecular trait, and for molecular trait on cancer risk in a direction consistent with the effect of the adiposity trait on cancer risk), we performed multivariable MR to estimate the mediating role of the molecular trait.

We found strong evidence that 13% of the causal effect of ASAT on increased endometrial cancer risk, overall and the endometrioid subtype, was mediated by lower SHBG, (95% CI = 5 to 20% and 5 to 21% for overall and endometrioid endometrial cancer, respectively), though we note that these analyses may suffer from weak instrument bias (as conditional F-statistics for ASAT were <10), which can bias effect estimates towards or away from the null in multivariable MR analyses[59] (**Table 5**, **Figure 5, Supplementary table 9**). There was also evidence for a mediating role of: higher genetically predicted bioavailable testosterone in the effect of ASAT on increased risks of endometrial cancer (percent mediated = 11, 95% CI = 2 to 20%) and endometrioid endometrial cancer (percent mediated = 11, 95% CI = 2 to 21%), though these analyses may also suffer from weak instrument bias; lower IGFBP-1 in the effect of ASAT on increased oesophageal adenocarcinoma risk (percent mediated = 42, 95% CI = 7 to 77%); higher genetically predicted adiponectin in the effect of GFAT on reduced non-endometrioid endometrial cancer risk (percent mediated = 27, 95% CI = 1 to 52%); and higher genetically predicted fasting insulin in the effect of ASAT on increased risks of endometrial cancer (percent mediated = 35, 95% CI = 1 to 68.1%) and endometrioid endometrial cancer (percent mediated = 40, 95% CI = 0 to 80%) (**Table 5**, **Figure 5, Supplementary table 10**). The resulting relationships between adiposity distribution traits, molecular traits, and cancer risks based on our univariable and multivariable MR analyses are shown in **Figures 6 and 7**. Note that conditional F-statistics were also < 10 for multivariable MR analyses examining a potential mediating role of CXCL-8 in the effect of ASAT on ovarian cancer risk, and HDL cholesterol in the effect of ASAT on triple negative or basal-like breast cancer risk, meaning we may be underestimating the mediating role of these traits in these relationships.

**Figure 5.**
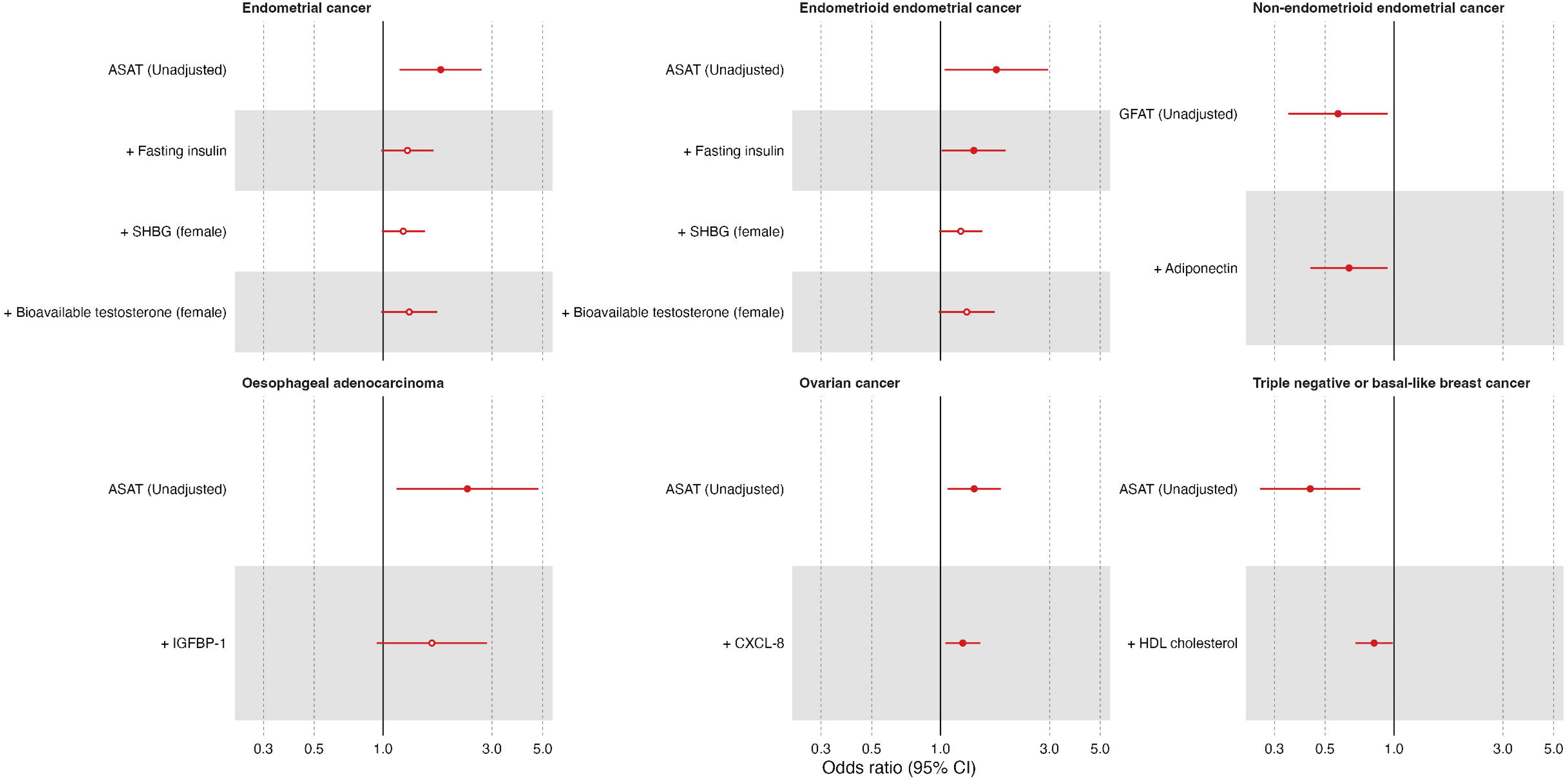
Univariable and multivariable estimates of measures of adiposity on risks of obesity-related cancers, adjusting sequentially for each possible mediator. ASAT = adipose subcutaneous adipose tissue; GFAT = gluteofemoral adipose tissue; SHBG = sex hormone-binding globulin; IGFBP = IGF binding protein; CXCL = C-X-C motif chemokine ligand; HDL = high-density lipoprotein.

**Figure 6.**
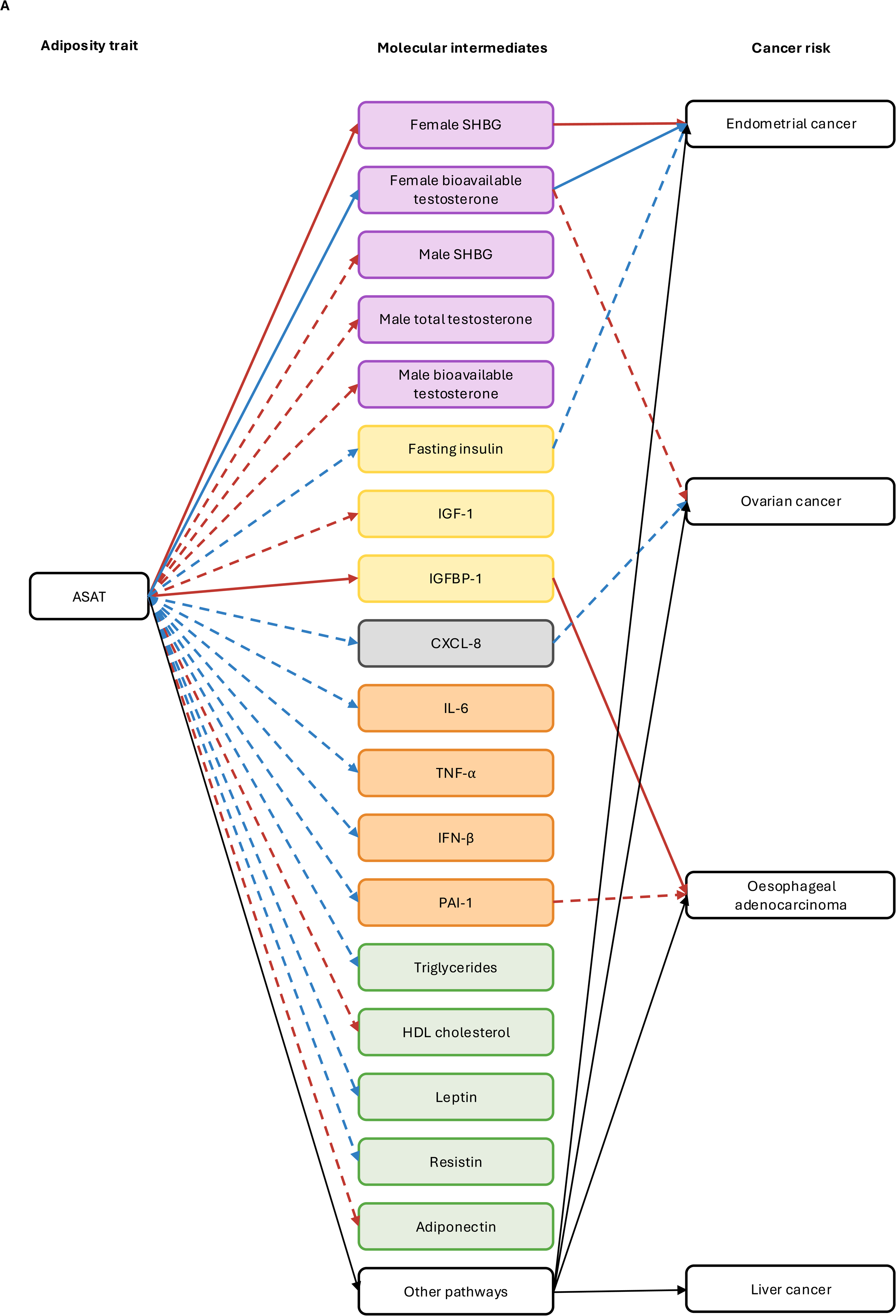
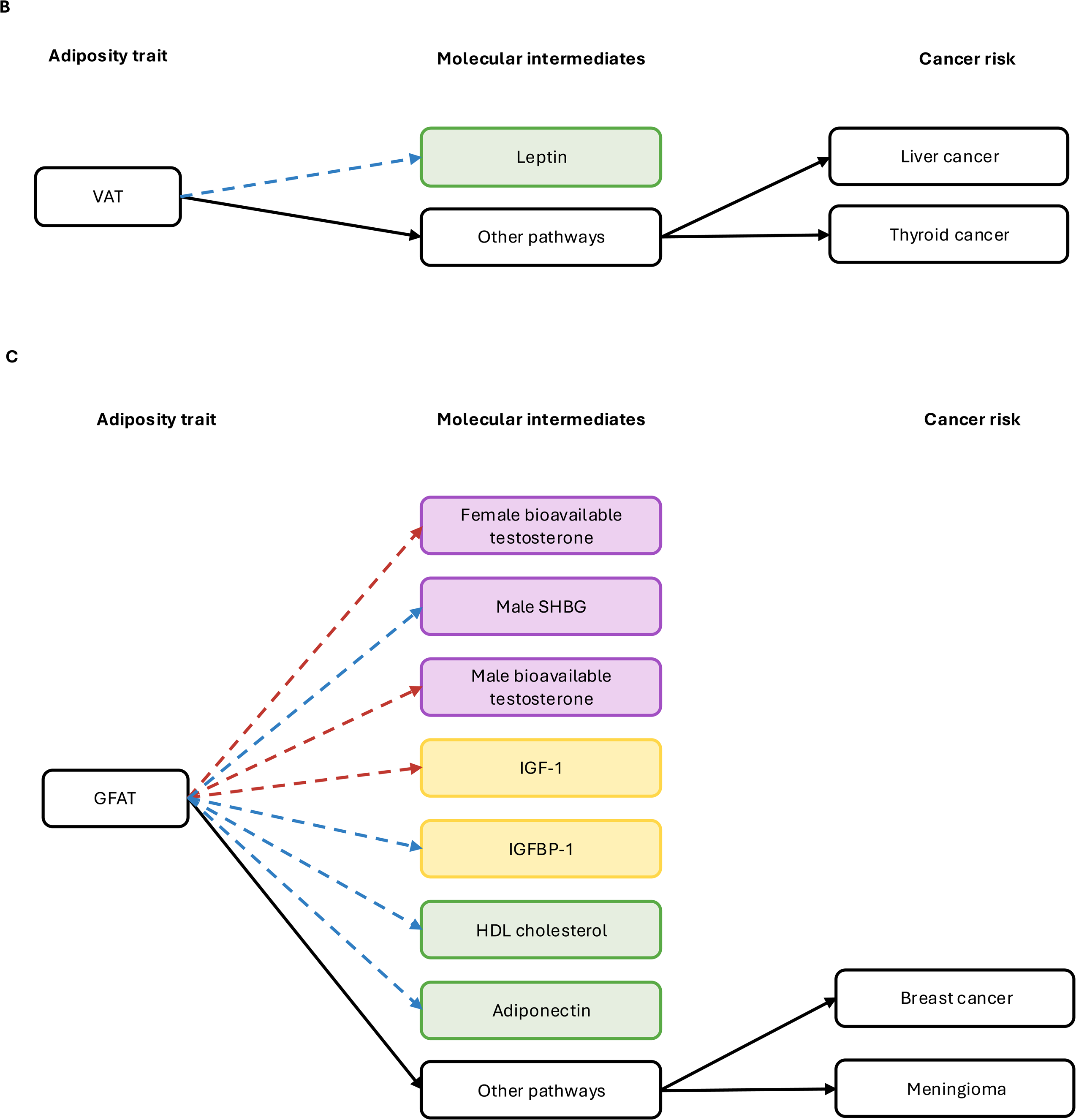
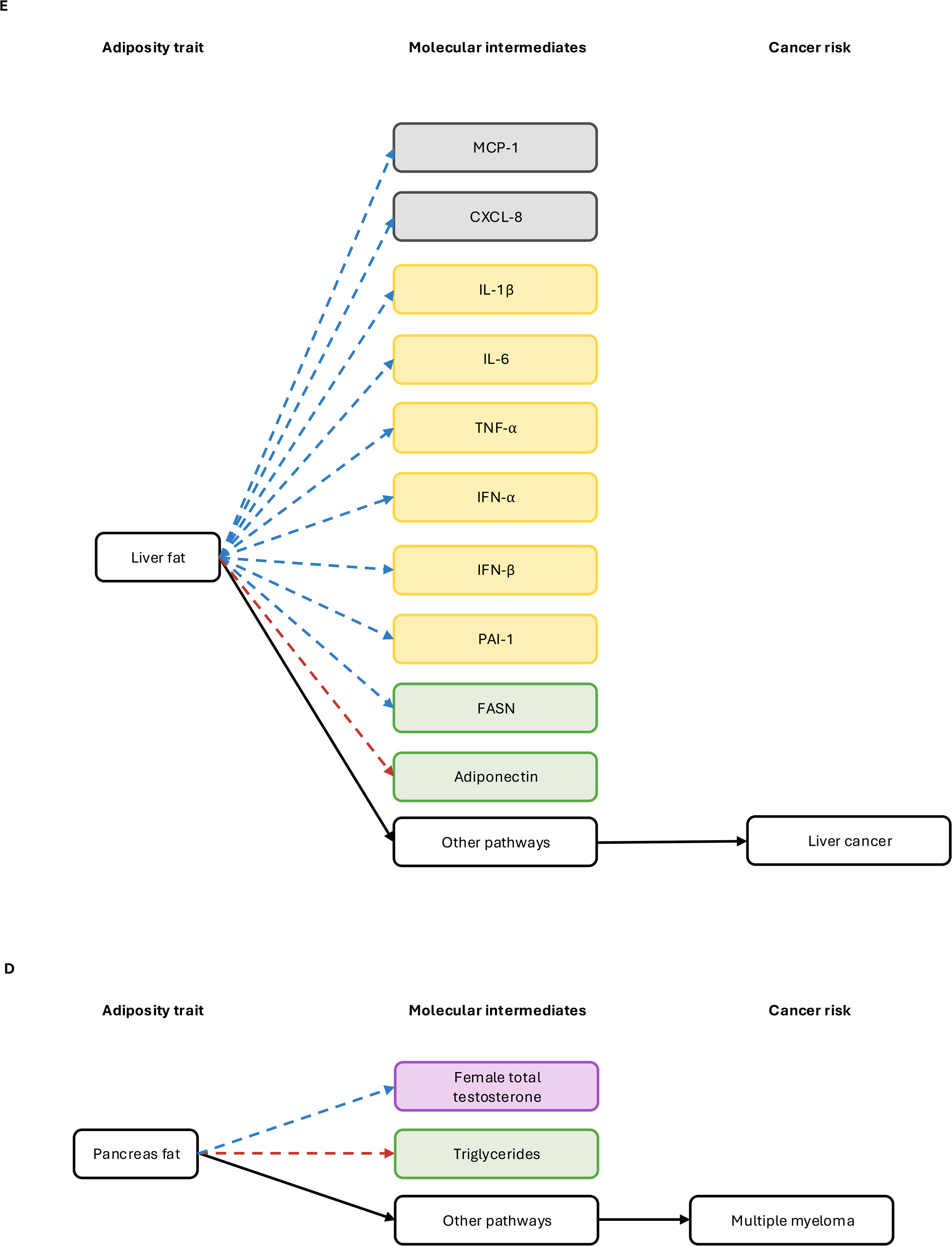
Schematic showing the results from MR analyses of adiposity measures on molecular traits, and molecular traits on cancer risks (overall only, i.e. not including cancer subtypes/subsites). Purple molecular traits are sex hormones and related traits; yellow molecular traits are insulin-related traits; orange molecular traits are inflammation-related adipokines; green molecular traits are lipid-related traits; grey traits are chemokine traits. Red arrows represent analyses with evidence for a causal effect which increases molecular trait levels or cancer risk; blue arrows represent analyses with evidence for a causal effect which decreases molecular trait levels or cancer risk. Solid arrows represent analyses for which there was evidence for a mediating effect of the molecular trait in multivariable MR analyses; dotted arrows represent analyses where there was evidence in univariable MR analyses only. All black arrows represent biological pathways which were not evaluated as part of the analyses outlined in this manuscript, but are presumed to exist given no molecular trait evaluated was estimated to mediate 100% of the effects of adiposity measures on cancer risks. **A** MR results from analyses relating to ASAT. **B** MR results from analyses relating to VAT. **C** MR results from analyses relating to GFAT. **D** MR results from analyses relating to pancreas fat. **E** MR results from analyses relating to liver fat.

**Figure 7.**
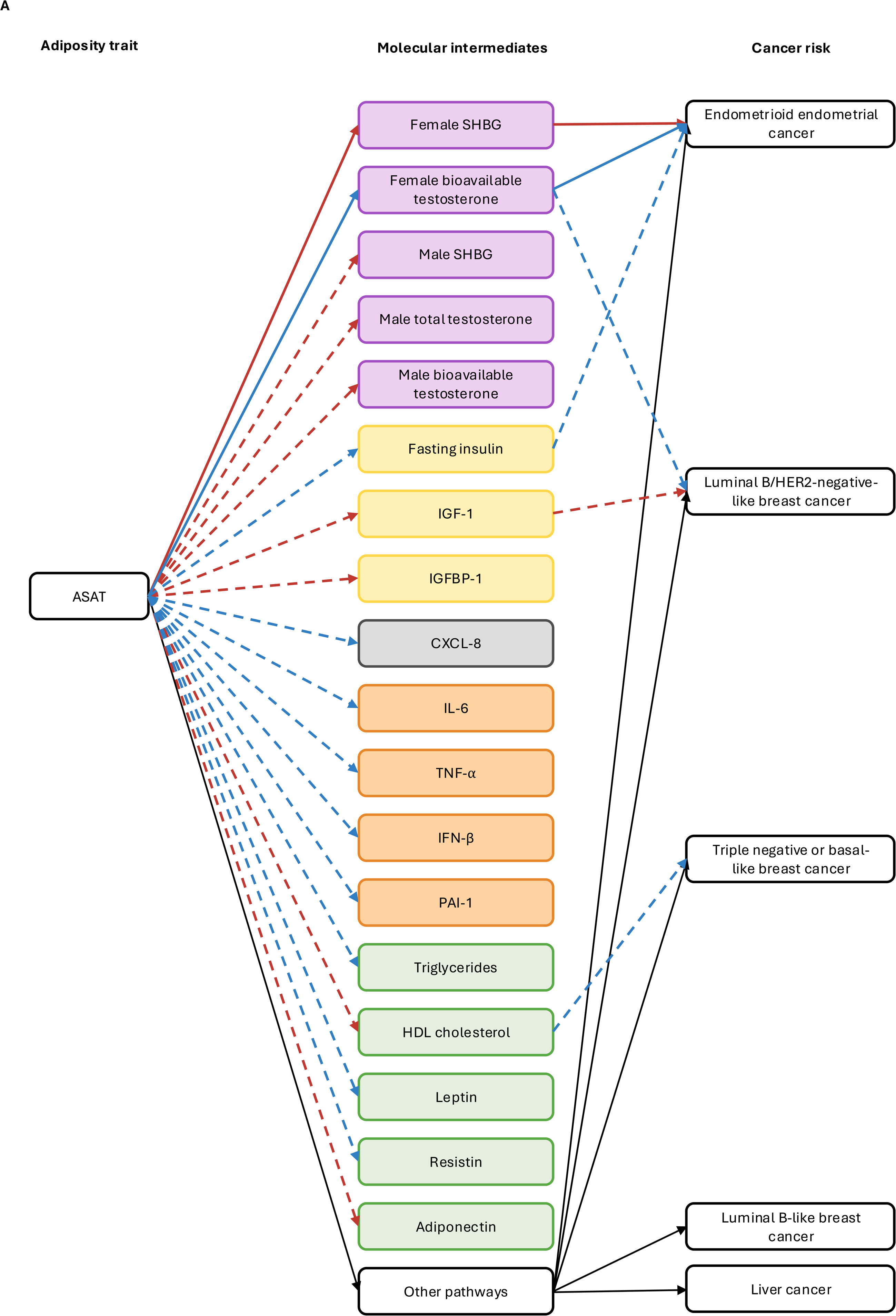
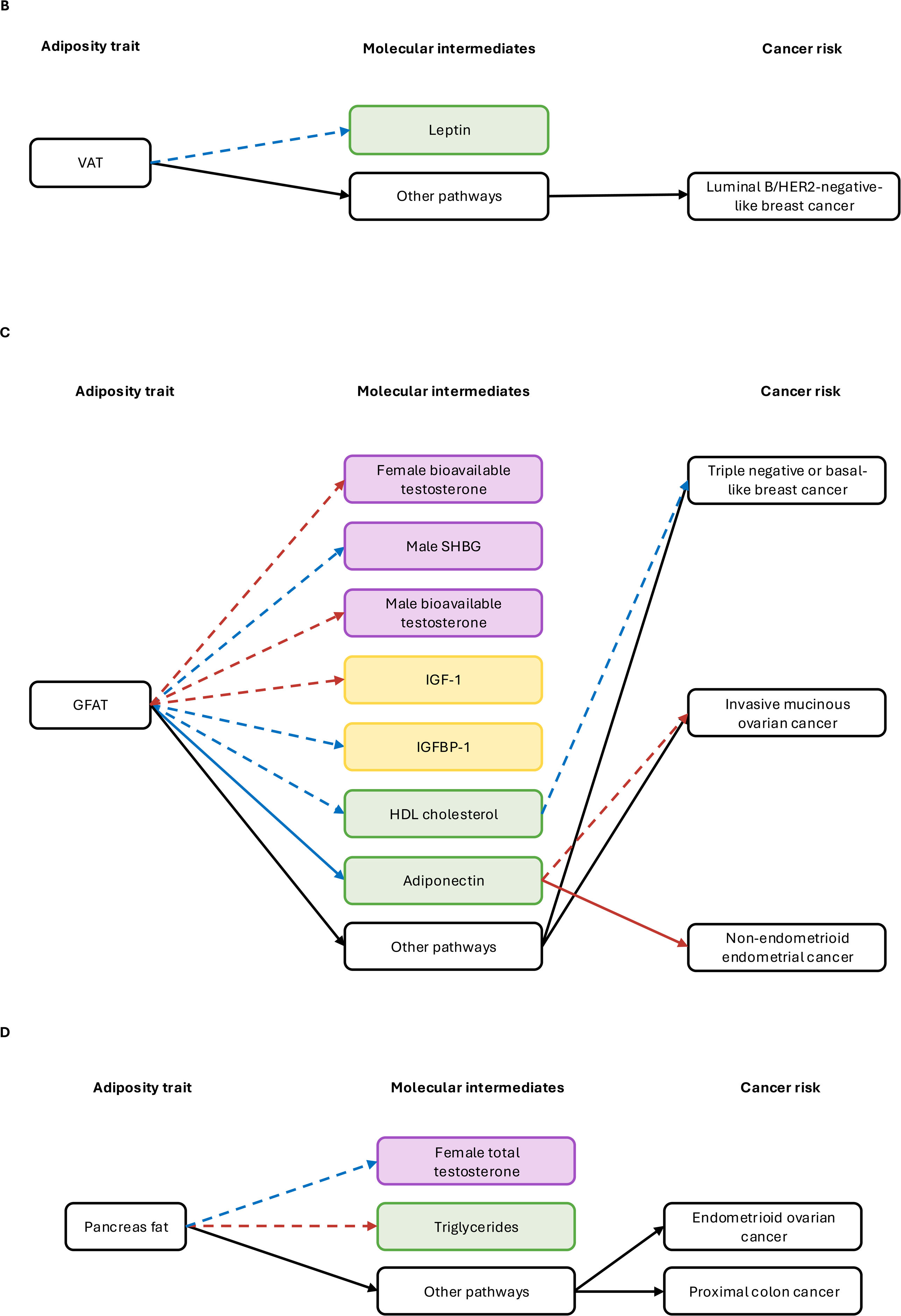
Schematic showing the results from MR analyses of adiposity measures on molecular traits, and molecular traits on subtype/subsite-specific cancer risks. Purple molecular traits are sex hormones and related traits; yellow molecular traits are insulin-related traits; orange molecular traits are inflammation-related adipokines; green molecular traits are lipid-related traits; grey traits are chemokine traits. Red arrows represent analyses with evidence for a causal effect which increases molecular trait levels or cancer risk; blue arrows represent analyses with evidence for a causal effect which decreases molecular trait levels or cancer risk. Solid arrows represent analyses for which there was evidence for a mediating effect of the molecular trait in multivariable MR analyses; dotted arrows represent analyses where there was evidence in univariable MR analyses only. All black arrows represent biological pathways which were not evaluated as part of the analyses outlined in this manuscript, but are presumed to exist given no molecular trait evaluated was estimated to mediate 100% of the effects of adiposity measures on cancer risks. **A** MR results from analyses relating to ASAT. **B** MR results from analyses relating to VAT. **C** MR results from analyses relating to GFAT. **D** MR results from analyses relating to pancreas fat.

There was potential sample overlap (which can compound weak instrument bias) in multivariable MR analyses evaluating evidence for a causal effect of ASAT adjusted for HDL cholesterol, SHBG, and bioavailable testosterone on cancer risks. Sensitivity analyses examining the impact of sample overlap on multivariable MR analyses involving HDL cholesterol revealed consistent results with those obtained from primary analyses (**Supplementary tables 10 and 11, Supplementary figure 6**). Due to the lack of alternative female-specific summary genetic data for SHBG and bioavailable testosterone, sensitivity analyses examining the impact of sample overlap on these traits could not be performed. Given the assumption of a covariance of 0 is unlikely to be the case where sample overlap exists, we anticipate we may be underestimating weak instrument bias for multivariable MR analyses including these traits.

## Discussion

We conducted two-sample and multivariable MR analyses using the largest available GWAS that included ∼40,000 individuals of European ancestry to investigate the role of five understudied adiposity traits in the risks of 12 obesity-related cancers, and cancer subtypes/subsites. We found evidence for a causal effect of higher genetically predicted liver fat on increased liver cancer risk, higher genetically predicted pancreas fat on increased endometrioid ovarian cancer risk, and higher genetically predicted ASAT on decreased luminal B/HER2-negative-like and triple negative or basal-like breast cancer risk. We also found evidence for a causal effect of adiposity traits on several more cancer types, with variable directions of effect, and ASAT showing the most consistent evidence for an effect on cancer types, with evidence for a causal risk-increasing effect on four of the 12 overall cancer types investigated. We found little evidence for a causal effect of the other adiposity distribution traits investigated, except for liver fat and VAT on liver cancer risk and a protective effect of GFAT on breast and ovarian cancer risks. Subsequent analyses highlighted a potentially mediating role of SHBG in the effect of ASAT on endometrial cancer (overall and the endometrioid subtype), though we note the potential for weak instrument bias in these analyses. We also found evidence for a mediating role of bioavailable testosterone in the effect of ASAT on risks of endometrial and endometrioid endometrial cancer, though these analyses may also suffer from weak instrument bias, IGFBP-1 in the effect of ASAT on oesophageal adenocarcinoma, adiponectin in the effect of GFAT on non-endometrioid endometrial cancer, and fasting insulin and bioavailable testosterone in the effect of ASAT on risks of endometrial cancer and endometrioid endometrial cancer.

The importance of adiposity distribution (alongside the importance of total adiposity) in health outcomes is becoming increasingly recognised. This has been mostly driven by previous research on cardiovascular outcomes, which has highlighted seemingly harmful effects of VAT, ASAT, liver fat and pancreas fat, but protective effects of GFAT[21,24,25]. However, whether the same causal effects of adiposity distribution on cancer risk is unclear. Our analysis suggests, for the first time, that this relationship is not as straightforward for cancer outcomes, with causal effects varying for the different measures of adiposity by cancer type. For instance, though we found evidence that higher genetically predicted pancreas fat increases risk of endometrioid ovarian cancer, we found evidence for a protective effect on proximal colon cancer. Similarly, we found evidence for a protective effect of higher genetically predicted ASAT on risks of luminal B/HER2-negative-like and triple negative or basal-like breast cancer, yet there was evidence for increased risk of endometrial, liver, oesophageal adenocarcinoma, and ovarian cancer. Evidence for an effect was also seen for liver fat and liver cancer risk. An association between central adiposity (including liver fat) and liver cancer is well-established in previous conventional observational analyses[60,61]. In the multivariable MR analysis conducted to assess whether the genetic instruments for ASAT and liver fat were capturing shared mechanistic pathways, the effect estimates for both traits were attenuated towards the null compared with the univariable MR analyses. This suggests that the instruments for ASAT and liver fat may reflect overlapping biological mechanisms, potentially related to central adiposity more broadly. This interpretation is supported by the univariable MR analyses, which also identified an effect of VAT on liver cancer risk. By contrast, there was little evidence for a causal effect of BMI on liver cancer risk. This underscores the importance of fat distribution, rather than overall body size, in influencing certain outcomes. Overall, our results show that the effects of adiposity distribution traits on cancer outcomes appear likely to vary, both by adiposity trait and cancer type.

As well as evidence for differential effects of adiposity traits across cancer sites, we also found evidence for variable effects on molecular traits. For instance, we found evidence that both increased ASAT and GFAT reduced IGF-1 levels. However, we found evidence that ASAT and GFAT had opposing effects on IGFBP-1 levels. Similarly differential effects of the adiposity traits are also seen for the sex hormones, chemokines, fatty acid-related traits, and adipokines investigated. The different adipose depots are known to vary based on the makeup of cells, and thus, it may not be surprising that changes to the volume of adiposity at the different sites results in dysregulation of different pathways[16]. These complex molecular interactions may explain the variable effects on cancer risk seen for each adiposity trait.

We found evidence suggesting a mediating role of SHBG, fasting insulin, bioavailable testosterone, and IGFBP-1 in the effects of ASAT on obesity-related cancers. Our analysis identified a potentially mediating role of SHBG in the effect of ASAT on endometrial cancer (overall and the endometrioid subtype) risk, and for a mediating role of bioavailable testosterone and fasting insulin in this relationship though it should be noted that these analyses suffer from weak instruments and effects may therefore be biased in either direction so should be interpreted with caution[59]. These results are similar to those for BMI obtained previously[19,44]. Should these results be replicated in future analyses which avoid weak instrument bias, this would fit with the “unopposed oestrogen” hypothesis for the development of endometrial cancer and the endometrioid subtype, which postulates that endometrial cancer develops as a result of increased oestrogen (which is influenced by insulin levels and is a direct product of the aromatization of testosterone) unopposed by SHBG[44].

We also found evidence suggesting a mediating role of adiponectin in the effect of GFAT on the risk of non-endometrioid endometrial cancer. In contrast to BMI, ASAT and liver fat, we found that higher genetically predicted GFAT increases adiponectin levels, which then may protect from non-endometrioid endometrial cancer development, thus in part explaining a potential protective effect of GFAT on non-endometrioid endometrial cancer risk.

Adiponectin is an inflammation-related adipokine produced by adipocytes, with a role in insulin sensitivity and fatty acid metabolism[62–65]. Our finding that higher genetically predicted GFAT increases adiponectin levels reflects previous observational analyses[66–68,68–78] and reflects the inflammatory profile of the genetic instruments used to proxy GFAT as characterised previously[21]. Adiponectin is widely recognised as being important for maintaining cardiometabolic health and is also associated with decreased risk of endometrial cancer in observational analyses[79–84]. Our findings that the protective effect of GFAT on risk of non-endometrioid endometrial cancer may be partly explained by a favourable adiponectin profile fit with these previous analyses. Non-endometrioid endometrial cancers represent a minority of endometrial cancers, and little is known about their aetiology. Compared with endometrioid endometrial cancers, which have a very well-established link with obesity, previous evidence for an association between adiposity and non-endometrioid endometrial cancers is less clear. Our results suggest that a plausible reason for this could be that a harmful effect of overall adiposity may be conflated with a protective effect of higher adiposity specifically within GFAT, possibly explaining previous inconclusive results and demonstrating the importance of considering causal effects of distinct adipose deposits[44,85–90].

Finally, our analysis suggested that IGFBP-1 may mediate the effect of ASAT on oesophageal adenocarcinoma. The leading hypothesis for how obesity may cause oesophageal adenocarcinoma is through increased pressure, which can disrupt the gastroesophageal junction and increase risk of gastroesophageal reflux disease, a known risk factor for oesophageal adenocarcinoma[91,92]. Our finding that ASAT may have a causal effect on oesophageal adenocarcinoma fits with this hypothesis. However, we find novel evidence of IGFBP-1 having a potential molecular mediating role in the ASAT and oesophageal adenocarcinoma relationship. IGFBP-1 has a long-established inverse association with obesity, which is likely mediated by changes to insulin levels[93–95].

Consistent with this, the only measure of adiposity which had evidence for an effect on fasting insulin levels in our analysis was also ASAT. Previous observational research has highlighted an association between IGFBP-1 levels or genetic polymorphisms and oesophageal cancer risk[96–98]. The main physiological role of IGFBP-1 is thought to be the binding and regulation of IGFs, including IGF-1[99,100]. In our analysis, we found little evidence for a causal effect of IGF-1 directly on oesophageal adenocarcinoma risk, suggesting that the biological mechanism linking IGFBP-1 and oesophageal adenocarcinoma risk may be independent of its role regulating IGF-1. One possible explanation for this is through IGFBP-1 binding to integrin α5β1, which has a role in modulating cell motility, adhesion and proliferation, and has been linked previously to several cancers through its role in these processes[101,102]. Alternatively, IGFBP-1 may cause oesophageal adenocarcinoma through IGF-1 intercellular transport and intracellular activity, which may not be captured by circulating IGF-1 levels[101–108]. Thus, currently the relative importance of molecular traits such as IGFBP-1 remains uncertain in comparison with the mechanical effects of increased abdominal obesity in the development of oesophageal adenocarcinoma.

One important limitation of the analyses detailed here is that there is some overlap in the genetic instruments used as proxies for the different adiposity traits (**Supplementary table 2)**. However, no two adiposity traits share a complete set of SNPs, and where SNPs are shared the variant effect sizes and weightings used in the MR analyses also differ between traits, meaning that even where some SNPs are shared there is still a degree of specificity of genetic instruments. Supporting this, a major finding of our study is that the effect of adiposity on cancer risk seems to vary substantially depending on anatomical location of adipose depots, suggesting that genetic instruments for adiposity traits are capturing distinct biological signals. Future research should aim to further decipher the different signals being captured.

In terms of further limitations, our analyses were almost exclusively restricted to individuals of European ancestry, which limits the generalizability of our findings to other populations. Additionally, our analyses were limited by the availability of summary genetic data. For many cancer sites investigated in our analyses, there were no large-scale GWAS available, and thus these analyses likely lack power due to few cases being available in UK Biobank and FinnGen. Due to a lack of GWAS data in large, non-overlapping samples for traits, substantial sample overlap exists for several MR analyses described here. Though recent evidence suggests that this is unlikely to be a major issue in the absence of weak instrument bias, several of our multivariable MR analyses also suffered from conditionally weak instruments, potentially compounding this bias[31]. Similarly, although sex-specific sensitivity analyses were performed where data were available, these data were not available for all traits where there may be heterogeneity of instrument effects by sex.

Where data were available, in several of our sex-specific sensitivity analyses we found evidence for a causal effect where little evidence was seen in the primary analysis (for instance, we found evidence for an effect of female-specific but not sex-combined fasting insulin on risk of luminal B/HER2-negative-like breast cancer risk). This suggests that our use of genetic data derived from sex-combined GWAS for traits (other than sex hormones and SHBG) may be obscuring important biological mechanisms. Furthermore, we were unable to include two potentially mediating molecular traits, oestrogen and progesterone, in our analysis due to a lack of suitable genetic data. Several further traits in our analysis lacked suitable genetic instruments (as identified through our systematic approach), and therefore evidence for their causal effect on cancer outcomes could not be evaluated though they could be included as outcomes.

Another limitation of our MR analyses is that for many, we did not have enough genetic instruments to perform pleiotropy-robust models of MR, meaning the potential for violation of one of the key assumptions of MR could not be assessed. Moreover, the MR models utilised in our analyses assume linear relationships between exposures and outcomes, which is unlikely to be the case for all relationships investigated. MVMR additionally assumes that there is no exposure-mediator interaction, which cannot currently be assessed using summary-level data[56]. Furthermore, the summary genetic data for the adiposity traits were obtained using a deep learning model, an approach that has been shown to possibly introduce bias in SNP-exposure estimates[109]. Finally, we investigated circulating molecular traits as potential mediators of the effects of measures of adiposity on cancer risk. However, this does not allow for the investigation of local effects of adipose depots on local intermediates, nor for effects on activity and transport as opposed to total amount of molecular traits. In addition, further molecular or non-molecular traits (e.g. mechanical effects in the case of oesophageal adenocarcinoma) may have a role in mediating the effects of measures of adiposity on cancer risk but were not included in this analysis.

A benefit of using summary level data is that it overcomes the need for all traits (i.e. adiposity distribution, molecular traits, and cancer risk) to be measured in the same individuals. Though under certain assumptions MR can evaluate evidence for a causal effect between traits, the relationship between adiposity distribution and cancer risk is complex, with some level of pleiotropy between the different adiposity traits, as well as between adiposity distribution and molecular traits. Therefore, using MR alone, it can be difficult to ascertain directions of effect and true estimations of magnitudes of effect. Therefore, future research warrants investigation of the putative causal relationships suggested here in high-quality prospective studies.

## Conclusion

This analysis provides valuable insight into the potential causal effects of different anatomical locations of adiposity on obesity-related cancer risk, and highlights some of the potential mechanisms which may explain these effects. We show that the relationship between adiposity distribution and cancer risk is complex, with variable effects depending on the location of the adipose deposit and cancer type. Though future research is required to fully understand these relationships, this has important clinical implications for the management of obesity in cancer prevention. We also found evidence of possible distinct molecular mechanisms explaining the effect of ASAT on the risks of three different obesity-related cancers – endometrial cancer (overall and the endometrioid subtype), oesophageal adenocarcinoma, and ovarian cancer, though we note that these analyses may be impacted by weak instrument bias and should be interpreted with caution. These results highlight the possible importance of the distribution of adiposity as a cancer risk factor. Moreover, our results suggest that evaluating changes in adipose tissue distribution may have a role in future obesity treatment and cancer prevention interventions.

## Supporting information

Figures high quality

## Data Availability

All data produced in the present study are available upon reasonable request to the authors

## Acknowledgements

EH is supported by a Cancer Research UK Population Research Committee Studentship (C18281/A30905), the CRUK Integrative Cancer Epidemiology Programme (C18281/A29019) and is part of the Medical Research Council Integrative Epidemiology Unit at the University of Bristol which is supported by the Medical Research Council (MC_UU_00032/03) and the University of Bristol. LV acknowledges support from Czech Health Research Council by project NU21-03-00145. LJG is supported by a Cancer Research UK 25 (C18281/A29019) programme grant (the Integrative Cancer Epidemiology Programme). NT is supported by Cancer Research UK (PRCPJT-May22\100028) and is also part of the Medical Research Council Integrative Epidemiology Unit at the University of Bristol which is supported by the Medical Research Council. EEV is supported by the World Cancer Research Fund (WCRF UK), as part of the World Cancer Research Fund International grant program (IIG_FULL_2024_029).

This work used the computational facilities of the Advanced Computing Research Centre, University of Bristol - http://www.bristol.ac.uk/acrc/.

The funders had no role in study design, data collection and analysis, decision to publish, or preparation of the manuscript.

We want to acknowledge the participants and investigators of the FinnGen study. The breast cancer genome-wide association analyses were supported by the Government of Canada through Genome Canada and the Canadian Institutes of Health Research, the ‘Ministère de l’Économie, de la Science et de l’Innovation du Québec’ through Genome Québec and grant PSR-SIIRI-701, The National Institutes of Health (U19 CA148065, X01HG007492), Cancer Research UK (C1287/A10118, C1287/A16563, C1287/A10710) and The European Union (HEALTH-F2-2009-223175 and H2020 633784 and 634935). All studies and funders are listed in Michailidou et al (Nature, 2017). Data on glycaemic traits have been contributed by MAGIC investigators and have been downloaded from www.magicinvestigators.org.

The authors would like to thank the participants of the many studies from which data were used in this analysis, including: GECCO, CCFR, CORECT, BCAC, ECAC, OCAC, UK Biobank, FinnGen, Twin Study, MAGIC, DECODE, BEACON.

## Statistical analyses and data availability

Statistical analyses were performed using R (Vienna, Austria)[110] version 4.0.2. Univariable MR analyses were performed using “TwoSampleMR”[52,111] (version 0.5.6) and MVMR analysis with “MVMR”[59] (version 0.3). Proxy SNPs were identified using “LDlinkR”[112] (version 1.2.3) and LD reference panels were compiled using “ieugwasr”[113] (version 0.1.5). R packages “gwasvcf”[114] (version 0.1.1), “gwasglue”[115] (version 0.0.0.9000), “VariantAnnotation”[116] (version 1.36.0), and “remotes”[117] (version 2.4.2) were also used for some MR analyses. R packages “ggforestplot”[118] (version 0.1.0) and “ggplot2”[119] (version 3.4.2) were used to create the plots used in figures. Some GWAS data were accessed through the OpenGWAS database API[111,120] (see **Table 1**; accessed on: 01/01/2024). METAL[38] (version 2011-03-25) was used for the GWAS meta-analyses. All scripts used to carry out this analysis are available at: https://github.com/EmmaHazelwood/Adiposity-distribution-cancer-risk-MR.git. Power calculations were performed using https://sb452.shinyapps.io/power/. All GWAS data generated are available at: (link to be added).

## Disclaimer

Where authors are identified as personnel of the International Agency for Research on Cancer/ World Health Organization, the authors alone are responsible for the views expressed in this article and they do not necessarily represent the decisions, policy or views of the International Agency for Research on Cancer/ World Health Organization.

## Tables

**Table 1.** Details of GWAS used in MR analyses. GWAS = genome-wide association study, MR = Mendelian randomization, BMI = body mass index, SHBG = sex hormone-binding globulin, HDL = high-density lipoprotein.

**Table 2.** Results of MR analyses examining the effect of measures of adiposity on risk of obesity-related cancers.

**Table 3.** Results of MR analyses examining the effect of measures of adiposity on potential molecular mediators of the effect of adiposity on cancer risk.

**Table 4.** Results of MR analyses examining the effect of potential molecular mediators of the effect of adiposity distribution on cancer risk

**Table 5.** Results of multivariable MR mediation analyses.

## Supporting information

**Supplementary table 1.** Details of cancer cases and controls from UK Biobank and FinnGen which were meta-analysed.

**Supplementary table 2.** Genetic instruments used for each trait in MR analyses.

**Supplementary table 3.** Conditional F-statistics in multivariable MR combining all five adiposity distribution traits.

**Supplementary table 4** Results of MR analysis evaluating evidence for a causal effect of BMI on risks of obesity-related cancers.

**Supplementary table 5.** Results of sensitivity analyses examining the effect of sample overlap in MR analyses of adiposity traits and cancer-related molecular traits.

**Supplementary table 6.** Results of sex-specific MR analyses of adiposity trait, cancer-related molecular traits, and cancer risk.

**Supplementary table 7.** Conditional F-statistics in multivariable MR combining all all pairwise adiposity distribution traits.

**Supplementary table 8.** Results of multivariable MR mediation analyses examining effects of ASAT and liver fat on liver cancer risk.

**Supplementary table 9.** Conditional F-statistics in multivariable MR analyses evaluating a mediating role of molecular traits in the adiposity distribution-cancer risk relationship.

**Supplementary table 10.** Results of multivariable MR mediation analyses examining potential bias from sample overlap.

**Supplementary table 11.** Conditional F-statistics in multivariable MR analyses examining potential bias from sample overlap.

**Supplementary note.** MR-STROBE checklist.

## Supplementary methods

**Supplementary figure 1.** Genetic correlation between adiposity distribution traits. ASAT = adipose subcutaneous adipose tissue; VAT = visceral adipose tissue; GFAT = gluteofemoral adipose tissue; BMI = body mass index.

**Supplementary figure 2.** Results of power calculations for MR analyses estimating the causal effect of measures of adiposity on risk of obesity-related cancers. ASAT = adipose subcutaneous adipose tissue; VAT = visceral adipose tissue; GFAT = gluteofemoral adipose tissue.

**Supplementary figure 3.** Results of repeating univariable MR analyses without sample overlap. Odds ratios/betas shown are given as one SD increase in adiposity measure. Open/closed circles indicate the *P*-value did not/did meet the evidence threshold (*P*-value < 0.05) respectively. ASAT = adipose subcutaneous adipose tissue; VAT = visceral adipose tissue; GFAT = gluteofemoral adipose tissue; IGF = insulin-like growth factor; HDL = high-density lipoprotein.

**Supplementary figure 4.** Results of sex-specific univariable MR analyses of measures of adiposity on cancer risk; (A) overall and (B) by subtype. Odds ratios shown are given as one SD increase in adiposity measure. Open/closed circles indicate the *P*-value did not/did meet the evidence threshold (*P*-value < 0.05) respectively. ASAT = adipose subcutaneous adipose tissue; VAT = visceral adipose tissue; GFAT = gluteofemoral adipose tissue; BMI = body mass index.

**Supplementary figure 5.** Results of sex-specific univariable MR analyses of molecular traits on cancer risk. Open/closed circles indicate the *P*-value did not/did meet the evidence threshold (*P*-value < 0.05) respectively. ASAT = adipose subcutaneous adipose tissue; VAT = visceral adipose tissue; GFAT = gluteofemoral adipose tissue; SHBG = sex hormone-binding globulin; IGF = insulin-like growth factor; HDL = high-density lipoprotein.

**Supplementary figure 6.** Univariable and multivariable estimates of adiposity traits on risks of obesity-related cancers, adjusting sequentially for each possible mediator, to examine potential bias from sample overlap. Open/closed circles indicate the *P*-value did not/did meet the evidence threshold (*P*-value < 0.05) respectively. ASAT = adipose subcutaneous adipose tissue; HDL = high-density lipoprotein.

**Supplementary figure 7.** Univariable and multivariable estimates of ASAT and liver fat on risk of liver cancer. Open/closed circles indicate the *P*-value did not/did meet the evidence threshold (*P*-value < 0.05) respectively. ASAT = adipose subcutaneous adipose tissue.

