## Supplementary material for "Adiposity distribution and risks of twelve obesity-related cancers: a Mendelian randomization analysis": Supplementary methods.docx

**Study populations**

Summary genetic association data were obtained for breast cancer from a GWAS of 133,384 cases (including 63,767 luminal A-like; 15,942 luminal B/HER2-negative-like; 15,942 luminal B-like; 10,628 HER2-enriched-like; and 8,602 triple negative or basal-like breast cancer cases) and 113,789 controls (91,477 controls for subtype-specific GWAS) of European ancestry^1^. Summary genetic association data were obtained for colorectal cancer from a GWAS of 52,775 cases (including 28,736 colon; 14,416 proximal colon; 12,879 distal colon; and 14,150 rectal cancer cases) and 45,940 controls^2,3^. This sample excluded cases and controls from UK Biobank in order to avoid sample overlap with exposure datasets. Approximately 92% of participants in the overall colorectal cancer GWAS were of European ancestry (~8% were East Asian). All participants in the site-specific colorectal cancer GWAS were of European ancestry. Summary genetic association data were obtained for ovarian cancer from a GWAS of 25,509 cases (including 13,037 high grade serous carcinoma; 1,012 low grade serous carcinoma; 1,417 invasive mucinous; 2,810 endometrioid; 1,366 clear cell; and 3,103 low malignant potential ovarian cancer cases) and 40,941 controls of European ancestry^4^. Summary genetic association data were obtained for endometrial cancer from a GWAS of 12,906 cases (including 8,758 endometrioid and 1,230 non-endometrioid endometrial cancer cases) and up to 108,979 controls of European ancestry^5^. Summary genetic association data were obtained for oesophageal adenocarcinoma from a GWAS of 4,112 cases and 17,159 controls of European ancestry^6^.

Where data from large-scale cancer consortia and meta-analyses were not available, a meta-analysis was conducted using summary genetic data from UK Biobank and FinnGen, where available. This included: 2,085 cases and 635,902 controls for kidney (renal-cell) cancer^7,8^; 1,904 cases and 715,554 controls for thyroid cancer^8,9^; 1,836 cases and 715,344 controls for pancreas cancer^8,9^; 1,649 cases and 727,247 controls for multiple myeloma^8,9^; 862 cases and 715,717 controls for liver cancer^8,9^; and 279 cases and 715,718 controls for gallbladder cancer^8,9^. All participants included in the meta-analyses were of European ancestry. For further information on the numbers of cases and controls for UK Biobank and FinnGen separately see **Supplementary Table 1**. The full resulting summary genetic data from these meta-analyses is available for download from: (to be added). Summary genetic data were available from FinnGen only for meningioma, which included 1,012 cases and 259,583 controls of European ancestry^8^.

**Identification of potentially mediating molecular traits**

Where a generic term was used in the WCRF CUP report, e.g. “pro-inflammatory cytokines”, the cited literature was reviewed to determine suitable specific molecular traits to be included in the analysis. This approach identified five sex hormones or related traits (oestrogen, total testosterone^10^, bioavailable testosterone^10^, progesterone and sex hormone-binding globulin (SHBG))^7^, five insulin-related traits (fasting insulin^11^, insulin-like growth factor (IGF)-1^12^, IGF-2^13^, IGF binding protein (IGFBP)-1^13^, IGFBP-3^13^), seven pro-inflammatory cytokines (interleukin (IL)-1β^13^, IL-6^13^, tumour necrosis factor (TNF)-α^13^, C-reactive protein (CRP)^12^, interferon (IFN)-α^13^, IFN-β^13^, and plasminogen activator inhibitor (PAI)-1) ^13^, four inflammation-related adipokines (leptin^13^, visfatin^13^, resistin^13^, and adiponectin^13^), three lipid traits (triglycerides^14^, fatty acid synthase (FASN) ^13^, and high-density lipoprotein (HDL) cholesterol^14^, and two chemokines (monocyte chemotactic protein (MCP)-1^13^, and C-X-C motif chemokine ligand 8 (CXCL-8, also known as IL-8) ^13^).

**Mendelian randomization analyses**

For the primary MR analyses, an inverse variable weighted (IVW) random effects model was used where multiple genetic instruments were available. Where only a single genetic instrument was available, we calculated the Wald ratio to generate effect estimates. In order to assess the role of horizontal pleiotropy in biasing our results, where there were at least 10 SNPs used as genetic instruments, we re-calculated causal estimates using weighted median estimation and weighted mode estimation^15–19^. Each of these models makes different assumptions about horizontal pleiotropy, so comparing effect estimates to those obtained from the IVW method can provide complementary support in evaluating the potential of bias from horizontal pleiotropy (a violation of assumption (iii) of MR).

Where the effect allele frequency (EAF) was not reported in summary genetic data, or where EAF was considered intermediate (between 0.42 and 0.58), palindromic alleles were excluded from MR analyses. Where alleles with missing EAF were included (i.e. those with non-palindromic alleles so strand can be inferred), EAF was imputed based on the 1000 Genomes reference dataset to facilitate Steiger filtering. Where such SNPs were missing from the reference dataset (16 SNPs total, for MR analyses of IGF-1, resistin, fasting insulin, adiponectin, PAI-1, IGFBP-1, and CXCL-8 on risk of oesophageal adenocarcinoma), Steiger filtering could not be performed, meaning these SNPs were included without establishing whether they explain more variance in the exposure or outcome. Where SNPs used as genetic instruments were absent from the outcome dataset or excluded, proxy SNPs in high LD with the missing SNP (r^2^ > 0.8) were used where available.

**Sensitivity analyses**

We employed several sensitivity analyses to evaluate the reliability of our results. First, for all MR analyses where there was evidence for an effect (i.e. *P* < 0.05 in IVW or Wald ratio model and consistent direction of effect in further MR models if performed), and where analyses had substantial potential for sample overlap between the exposure and outcome GWAS, we repeated the analyses with alternative GWAS avoiding this overlap where data were available. Secondly, for any MR analyses where at least one of the traits is likely to have sex-specific genetic architecture (e.g. sex hormones and related traits), or where one GWAS was limited to a specific sex (e.g. breast and endometrial cancer), we repeated analyses with sex-specific GWAS where available, regardless of the strength of evidence in the sex-combined analyses. **Table** **1** shows further details on the GWAS used for these sensitivity analyses.
