## Supplementary material for "Adiposity distribution and risks of twelve obesity-related cancers: a Mendelian randomization analysis": Supplementary note.docx

**STROBE-MR checklist of recommended items to address in reports of Mendelian randomization studies**^1^ ^2^

| **Item No.** | **Section** | **Checklist item** | **Page No.** | **Relevant text from manuscript** |
| --- | --- | --- | --- | --- |
| 1 | **TITLE and ABSTRACT** | Indicate Mendelian randomization (MR) as the study’s design in the title and/or the abstract if that is a main purpose of the study |  | Adiposity distribution and risks of 12 obesity-related cancers: a Mendelian randomization analysis  We performed two-sample Mendelian randomization (MR) |
|  | **INTRODUCTION** |  |  |  |
| 2 | **Background** | Explain the scientific background and rationale for the reported study. What is the exposure? Is a potential causal relationship between exposure and outcome plausible? Justify why MR is a helpful method to address the study question | 1 | More specific measures, including abdominal subcutaneous, visceral, and gluteofemoral adipose tissue (ASAT, VAT, and GFAT, respectively), as well as the fat content of particular organs, in particular the liver and pancreas, have been shown to have differential roles in the effect of adiposity on risk of cardiometabolic outcomes^17,20^. However, the causal effect of these adiposity traits on cancer risk is less well understood. |
| 3 | **Objectives** | State specific objectives clearly, including pre-specified causal hypotheses (if any). State that MR is a method that, under specific assumptions, intends to estimate causal effects | 3 | Mendelian randomization (MR), a genetic epidemiological approach which uses genetic instruments as proxies to evaluate evidence for causal relationships between traits^21,22^. We used MR to evaluate the effects of five adiposity traits on risks of 12 obesity-related cancers and to evaluate the mediating effects of molecular features in the relationships between adiposity measures and obesity-related cancers. |
|  | **METHODS** |  |  |  |
| 4 | **Study design and data sources** | Present key elements of the study design early in the article. Consider including a table listing sources of data for all phases of the study. For each data source contributing to the analysis, describe the following: |  |  |
|  | a) | Setting: Describe the study design and the underlying population, if possible. Describe the setting, locations, and relevant dates, including periods of recruitment, exposure, follow-up, and data collection, when available. | 2 | Study populations section |
|  | b) | Participants: Give the eligibility criteria, and the sources and methods of selection of participants. Report the sample size, and whether any power or sample size calculations were carried out prior to the main analysis | 2 | Study populations section |
|  | c) | Describe measurement, quality control and selection of genetic variants | 3 | To construct genetic instruments for MR analyses, we obtained SNPs strongly (*P* < 5 × 10^−8^) and independently (*r*^2^ < 0.001) associated with each trait |
|  | d) | For each exposure, outcome, and other relevant variables, describe methods of assessment and diagnostic criteria for diseases | 2 | Study populations section |
|  | e) | Provide details of ethics committee approval and participant informed consent, if relevant | NA | NA |
| 5 | **Assumptions** | Explicitly state the three core IV assumptions for the main analysis (relevance, independence and exclusion restriction) as well assumptions for any additional or sensitivity analysis | 3 | MR can result in unbiased estimates of causal effects if the following assumptions are met: (i) the instrument strongly associates with the exposure, (ii) there is no confounding of the instrument-outcome relationship and (iii) the instrument only affects the outcome through the exposure^31^ |
| 6 | **Statistical methods: main analysis** | Describe statistical methods and statistics used |  |  |
|  | a) | Describe how quantitative variables were handled in the analyses (i.e., scale, units, model) |  | Table 1 |
|  | b) | Describe how genetic variants were handled in the analyses and, if applicable, how their weights were selected | NA | NA |
|  | c) | Describe the MR estimator (e.g. two-stage least squares, Wald ratio) and related statistics. Detail the included covariates and, in case of two-sample MR, whether the same covariate set was used for adjustment in the two samples | 4 | Where there was a single genetic instrument available for a trait, the Wald ratio and delta method were used to calculate effect estimates and approximate standard errors respectively^35^. Where there were multiple genetic instruments available, inverse-variance weighted (IVW) random-effects models were applied. Where there were more than 10 genetic instruments available for a trait, weighted median estimation and weighted mode estimation were used to evaluate evidence for horizontal pleiotropy^36–39^ |
|  | d) | Explain how missing data were addressed | NA | NA |
|  | e) | If applicable, indicate how multiple testing was addressed | 4 | As a heuristic, we applied a Bonferroni correction to account for multiple testing (*P* < 0.05/12 cancer types; 0.05/24 molecular traits; 0.05/18 unique adiposity-cancer pairs; or 0.05/10 multivariable MR analyses). *P*-values below this threshold were classed as “strong” evidence, whereas those between this threshold and 0.05 were classed as “suggestive” evidence. |
| 7 | **Assessment of assumptions** | Describe any methods or prior knowledge used to assess the assumptions or justify their validity | NA | NA |
| 8 | **Sensitivity analyses and additional analyses** | Describe any sensitivity analyses or additional analyses performed (e.g. comparison of effect estimates from different approaches, independent replication, bias analytic techniques, validation of instruments, simulations) | 4 | Where there were more than 10 genetic instruments available for a trait, weighted median estimation and weighted mode estimation were used to evaluate evidence for horizontal pleiotropy^36–39^  We employed several sensitivity analyses to evaluate the reliability of our results. These included repeating analyses while avoiding sample overlap and repeating MR analyses with a sex-specific outcome (e.g. breast and endometrial cancer) with sex-specific data where available. For more information on the MR analyses and sensitivity analyses conducted see **Supplementary methods**. |
| 9 | **Software and pre-registration** |  |  |  |
|  | a) | Name statistical software and package(s), including version and settings used | 4 | Statistical analyses were performed using R (Vienna, Austria) version 4.0.2. Univariable MR analyses were performed using “TwoSampleMR” (version 0.5.6) and MVMR analysis with “MVMR” (version 0.3). Proxy SNPs were identified using “LDlinkR” (version 1.2.3) and LD reference panels were compiled using “ieugwasr” (version 0.1.5). R packages “gwasvcf” (version 0.1.1), “gwasglue” (version 0.0.0.9000), “VariantAnnotation” (version 1.36.0), and “remotes” (version 2.4.2) were also used for some MR analyses. R packages “ggforestplot” (version 0.1.0) and “ggplot2” (version 3.4.2) were used to create the plots used in figures. Some GWAS data were accessed through the OpenGWAS database API (see **Table 1**; accessed on: 01/01/2024). METAL v2011-03-25 was used for the GWAS meta-analyses. All scripts used to carry out this analysis are available at: (add link). Power calculations were performed using <https://sb452.shinyapps.io/power/>. |
|  | b) | State whether the study protocol and details were pre-registered (as well as when and where) | NA | NA |
|  | **RESULTS** |  |  |  |
| 10 | **Descriptive data** |  |  |  |
|  | a) | Report the numbers of individuals at each stage of included studies and reasons for exclusion. Consider use of a flow diagram | NA | NA |
|  | b) | Report summary statistics for phenotypic exposure(s), outcome(s), and other relevant variables (e.g. means, SDs, proportions) | NA | NA |
|  | c) | If the data sources include meta-analyses of previous studies, provide the assessments of heterogeneity across these studies | 3 | Full meta-analysis results including heterogeneity P-value available to download |
|  | d) | For two-sample MR:  i.  Provide justification of the similarity of the genetic variant-exposure associations between the exposure and outcome samples  ii.  Provide information on the number of individuals who overlap between the exposure and outcome studies | 10 | European ancestries  Given the adiposity data were also generated using UK Biobank, it is likely that high levels of sample overlap are present for MR analyses using these data. |
| 11 | **Main results** |  |  |  |
|  | a) | Report the associations between genetic variant and exposure, and between genetic variant and outcome, preferably on an interpretable scale |  | Supplementary table 2 |
|  | b) | Report MR estimates of the relationship between exposure and outcome, and the measures of uncertainty from the MR analysis, on an interpretable scale, such as odds ratio or relative risk per SD difference |  | Tables 2-4 |
|  | c) | If relevant, consider translating estimates of relative risk into absolute risk for a meaningful time period | NA | NA |
|  | d) | Consider plots to visualize results (e.g. forest plot, scatterplot of associations between genetic variants and outcome versus between genetic variants and exposure) |  | Figures 2-4 |
| 12 | **Assessment of assumptions** |  |  |  |
|  | a) | Report the assessment of the validity of the assumptions | e.g. 4 | e.g. to avoid weak instrument bias (a violation of assumption (i) of MR) |
|  | b) | Report any additional statistics (e.g., assessments of heterogeneity across genetic variants, such as *I^2^*, Q statistic or E-value) | NA | NA |
| 13 | **Sensitivity analyses and additional analyses** |  |  |  |
|  | a) | Report any sensitivity analyses to assess the robustness of the main results to violations of the assumptions | 6-8 |  |
|  | b) | Report results from other sensitivity analyses or additional analyses | 6-8 | In sensitivity analyses, estimates derived from those examining the effect of sample overlap and sex-specific data were in a consistent direction with those derived from the primary analyses, although some 95% CIs crossed the null |
|  | c) | Report any assessment of direction of causal relationship (e.g., bidirectional MR) | NA | NA |
|  | d) | When relevant, report and compare with estimates from non-MR analyses | e.g. 9 | e.g. An association between central adiposity (including liver fat) and liver cancer is well-established in previous conventional observational analyses^49^ |
|  | e) | Consider additional plots to visualize results (e.g., leave-one-out analyses) | NA | NA |
|  | **DISCUSSION** |  |  |  |
| 14 | **Key results** | Summarize key results with reference to study objectives | 9 | understudied adiposity traits on risks of 12 obesity-related cancers. We found strong evidence for a causal effect of higher liver fat on liver cancer risk, higher pancreas fat on endometrioid ovarian cancer risk, and higher ASAT on luminal B/HER2-negative-like and triple negative or basal-like breast cancer risk. We also found suggestive evidence for a causal effect of adiposity traits on several more cancer types, with variable directions of effect. In subsequent analyses, we found evidence for a mediating role of SHBG in the effect of ASAT on endometrial cancer and in the effect of ASAT on endometrioid endometrial cancer. |
| 15 | **Limitations** | Discuss limitations of the study, taking into account the validity of the IV assumptions, other sources of potential bias, and imprecision. Discuss both direction and magnitude of any potential bias and any efforts to address them | 11 | However, several limitations exist. First… |
| 16 | **Interpretation** |  |  |  |
|  | a) | Meaning: Give a cautious overall interpretation of results in the context of their limitations and in comparison with other studies | 12 | We show that the relationship between adiposity distribution and cancer risk is complex, with variable effects depending on the location of the adipose deposit and cancer type. |
|  | b) | Mechanism: Discuss underlying biological mechanisms that could drive a potential causal relationship between the investigated exposure and the outcome, and whether the gene-environment equivalence assumption is reasonable. Use causal language carefully, clarifying that IV estimates may provide causal effects only under certain assumptions | e.g. 10 | e.g. This fits with the “unopposed oestrogen” hypothesis for the development of endometrial cancer and the endometrioid subtype |
|  | c) | Clinical relevance: Discuss whether the results have clinical or public policy relevance, and to what extent they inform effect sizes of possible interventions | 12 | our results suggest that evaluating changes in adipose tissue distribution in addition to a total reduction in adiposity could be an important aspect of future obesity treatment and cancer prevention interventions. |
| 17 | **Generalizability** | Discuss the generalizability of the study results (a) to other populations, (b) across other exposure periods/timings, and (c) across other levels of exposure | 11 | Our analysis was almost exclusively restricted to individuals of European ancestry, which limits the generalizability of our findings to other populations. |
|  | **OTHER INFORMATION** |  |  |  |
| 18 | **Funding** | Describe sources of funding and the role of funders in the present study and, if applicable, sources of funding for the databases and original study or studies on which the present study is based | 13 |  |
| 19 | **Data and data sharing** | Provide the data used to perform all analyses or report where and how the data can be accessed, and reference these sources in the article. Provide the statistical code needed to reproduce the results in the article, or report whether the code is publicly accessible and if so, where |  | All code and data available |
| 20 | **Conflicts of Interest** | All authors should declare all potential conflicts of interest | 13 |  |

This checklist is copyrighted by the Equator Network under the Creative Commons Attribution 3.0 Unported (CC BY 3.0) license.

1.	Skrivankova VW, Richmond RC, Woolf BAR, Yarmolinsky J, Davies NM, Swanson SA, et al. Strengthening the Reporting of Observational Studies in Epidemiology using Mendelian Randomization (STROBE-MR) Statement. JAMA. 2021;under review.

2.	Skrivankova VW, Richmond RC, Woolf BAR, Davies NM, Swanson SA, VanderWeele TJ, et al. Strengthening the Reporting of Observational Studies in Epidemiology using Mendelian Randomisation (STROBE-MR): Explanation and Elaboration. BMJ. 2021;375:n2233.
