## Supplementary figures and images for "Adiposity distribution and risks of twelve obesity-related cancers: a Mendelian randomization analysis"

### Figure 1.pdf

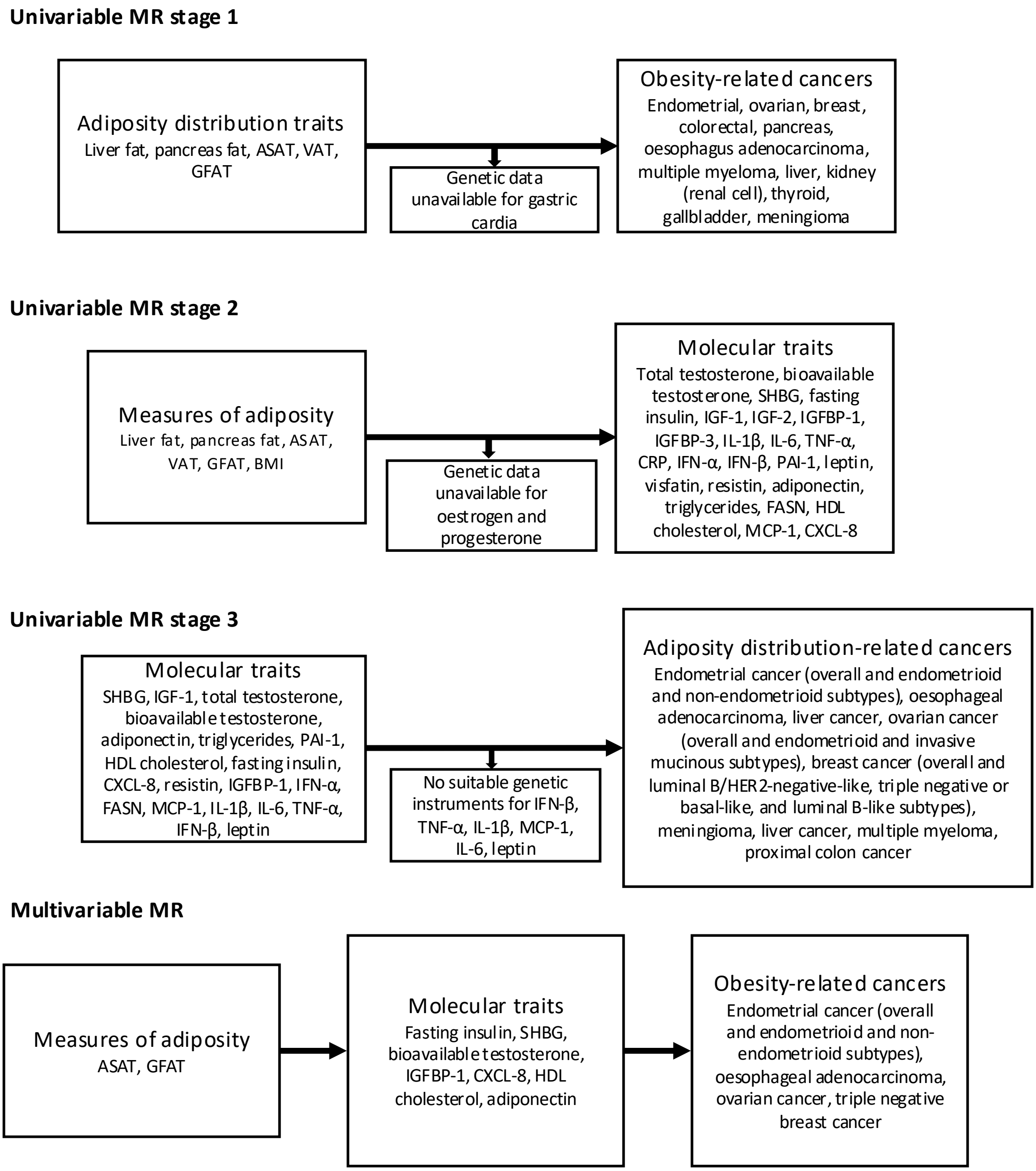

### Figure 2.png

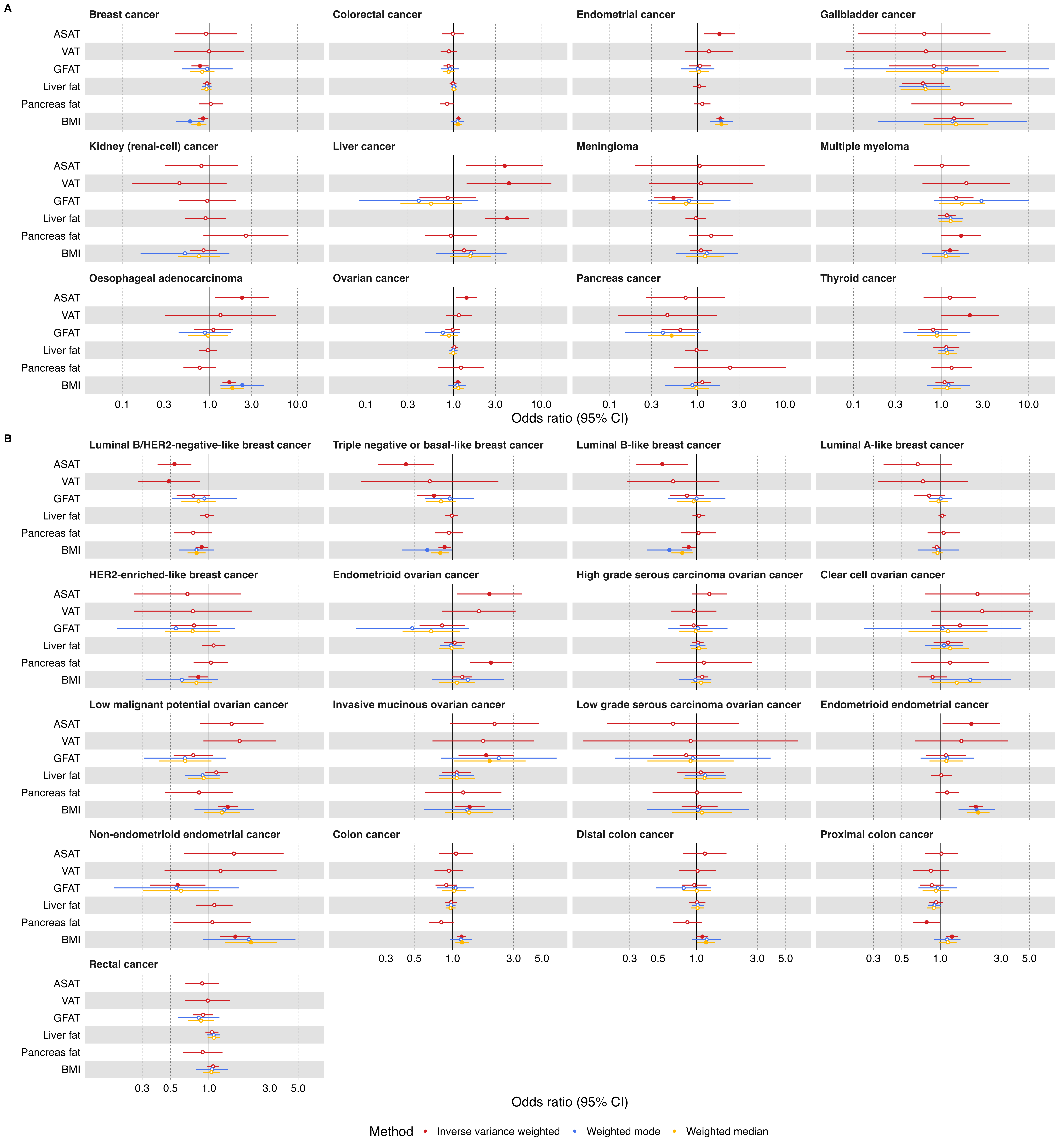

### Figure 3.png

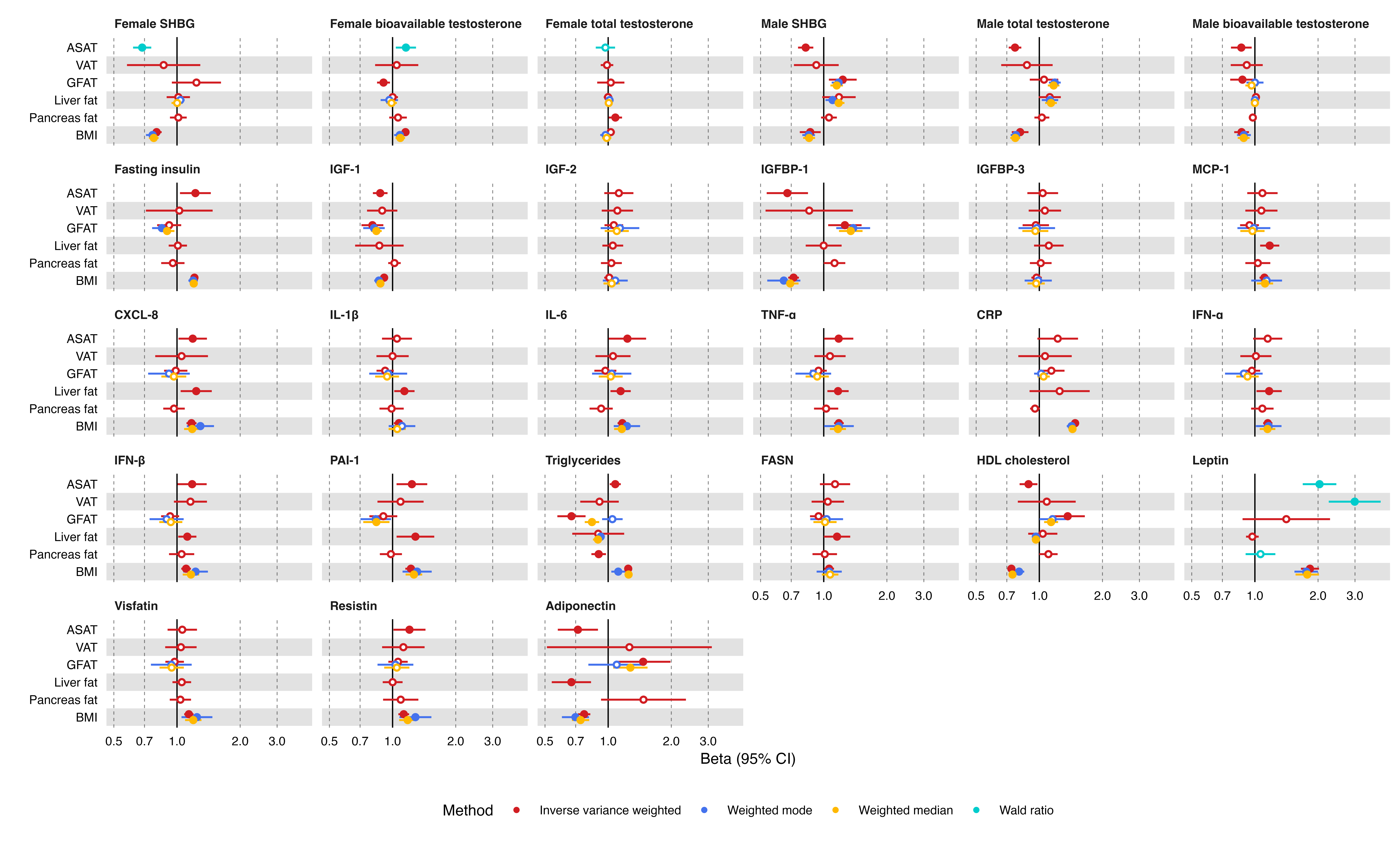

### Figure 4.png

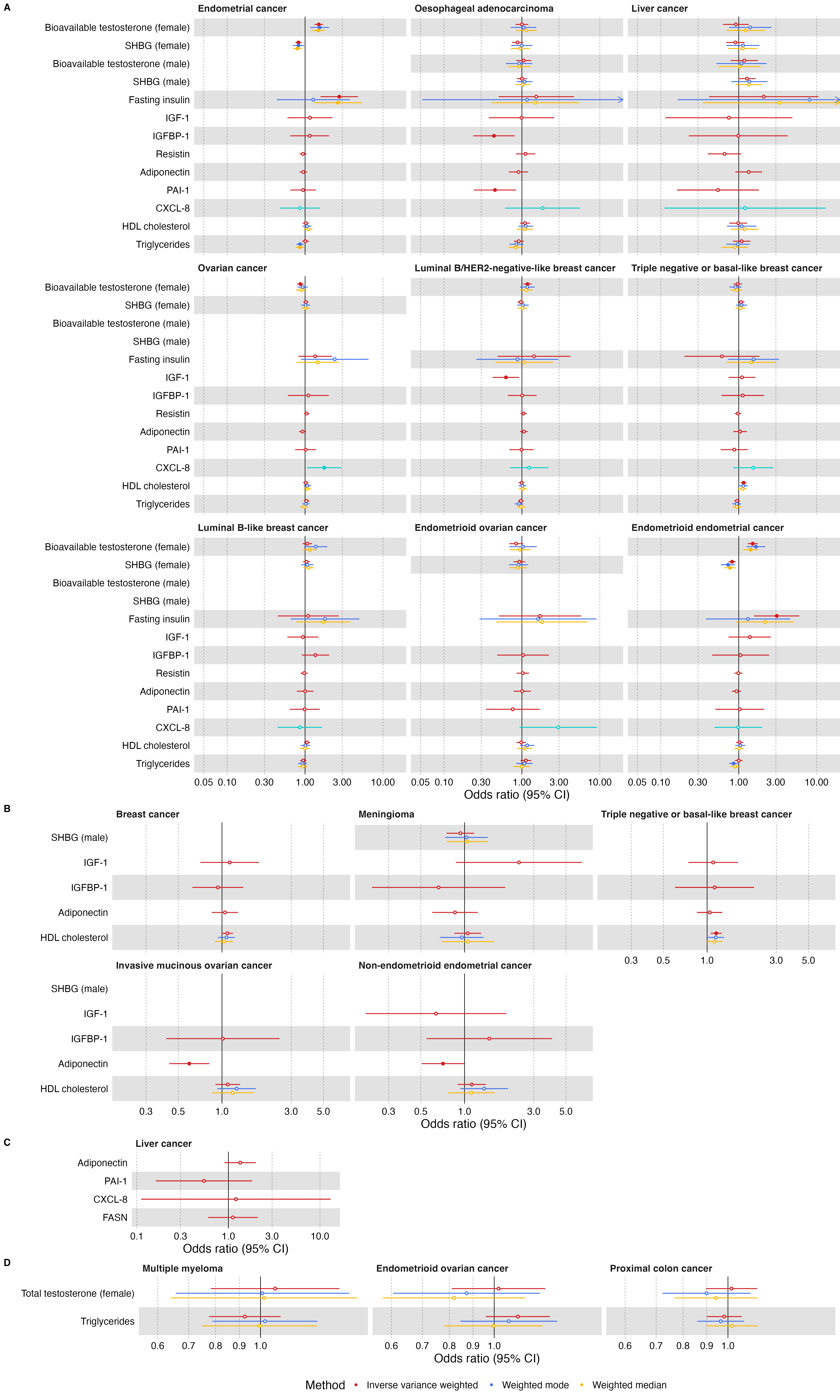

### Figure 5.png

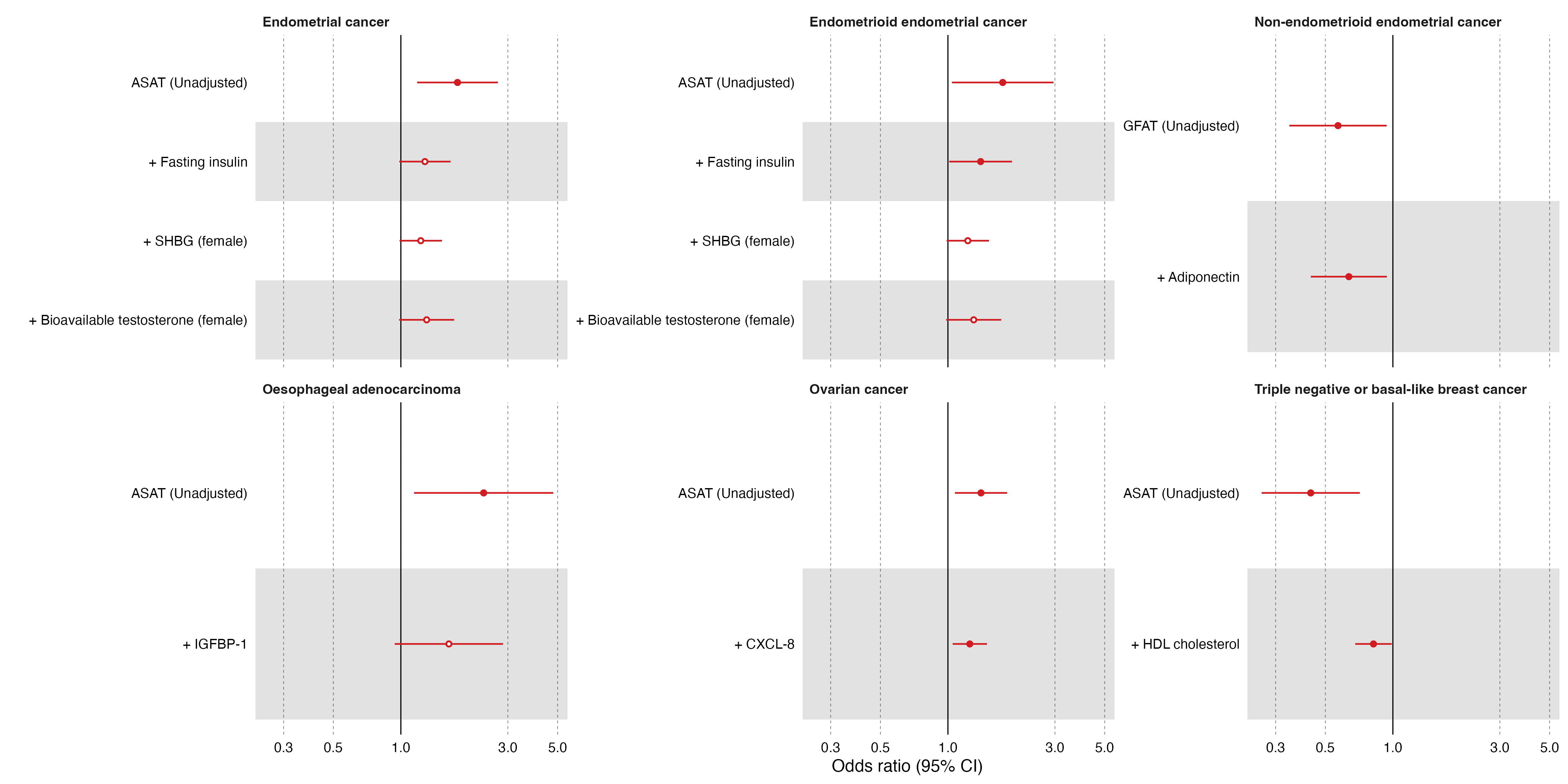

### Figures 6 and 7.pdf

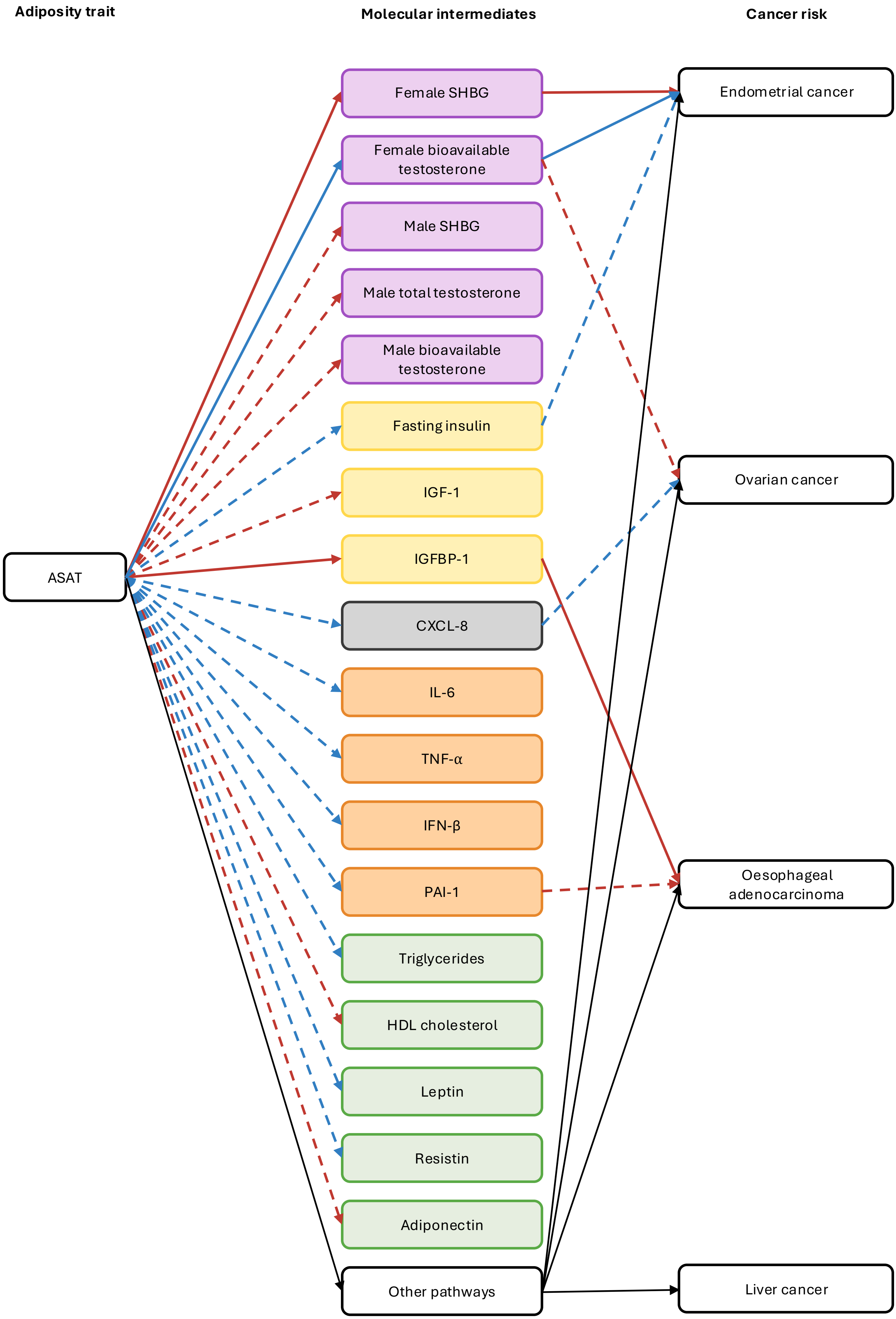

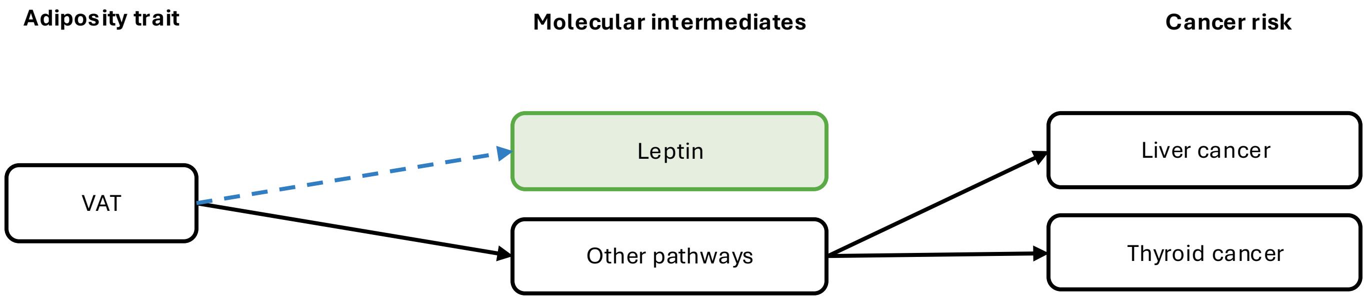

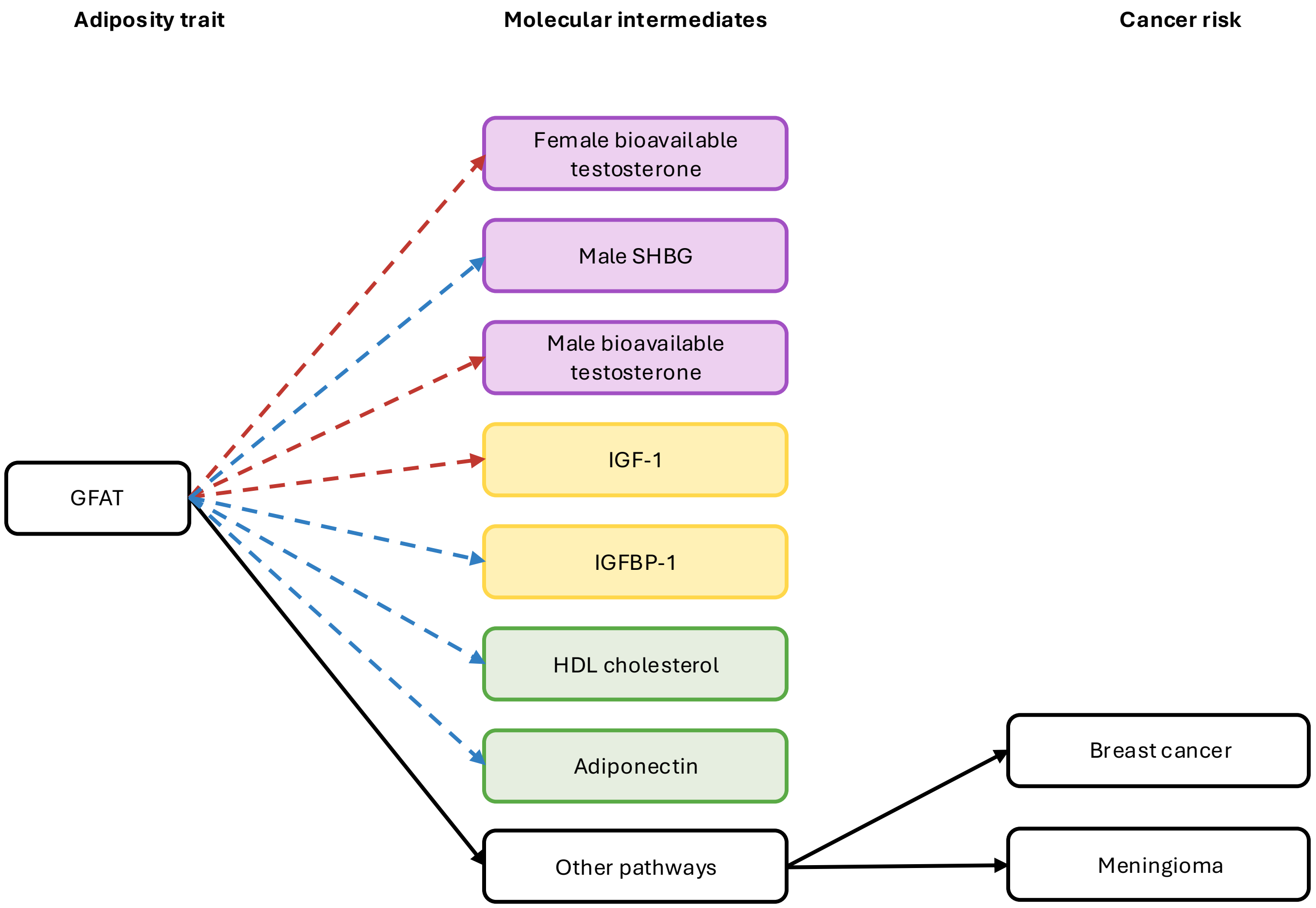

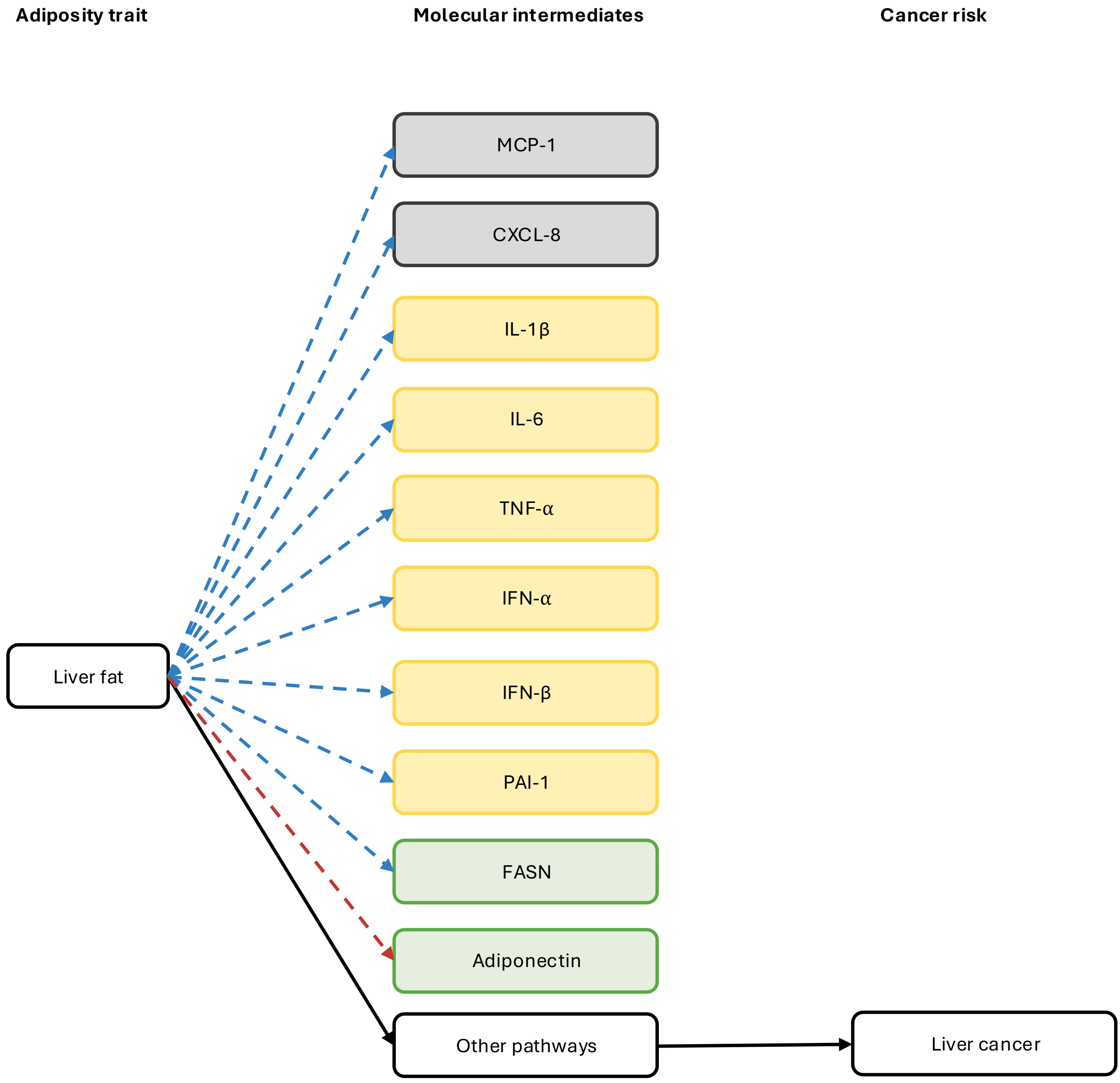

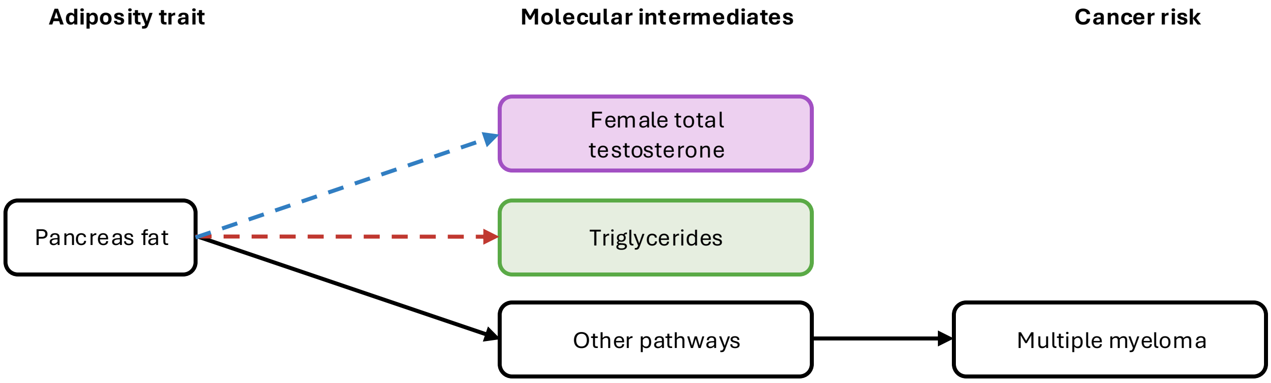

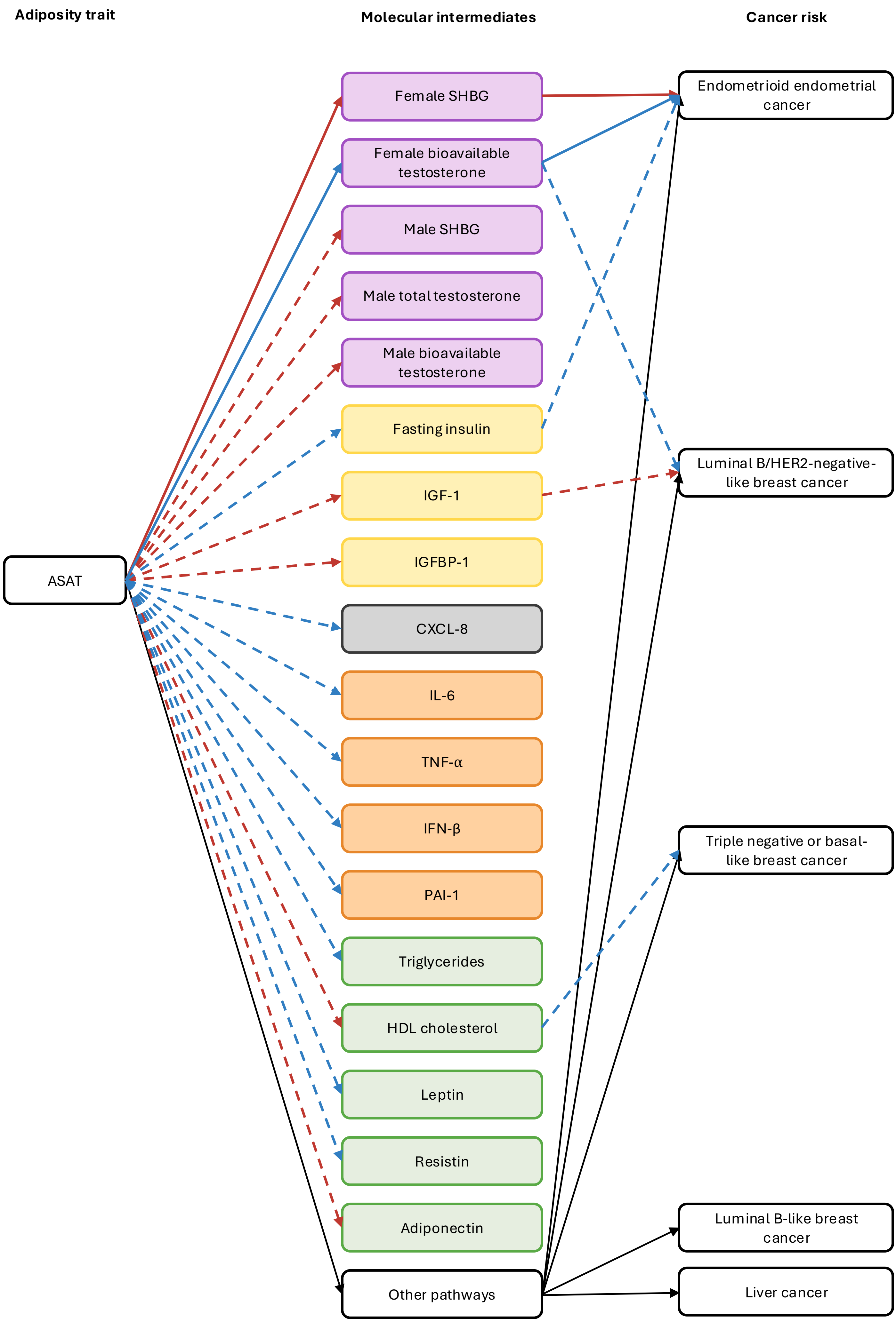

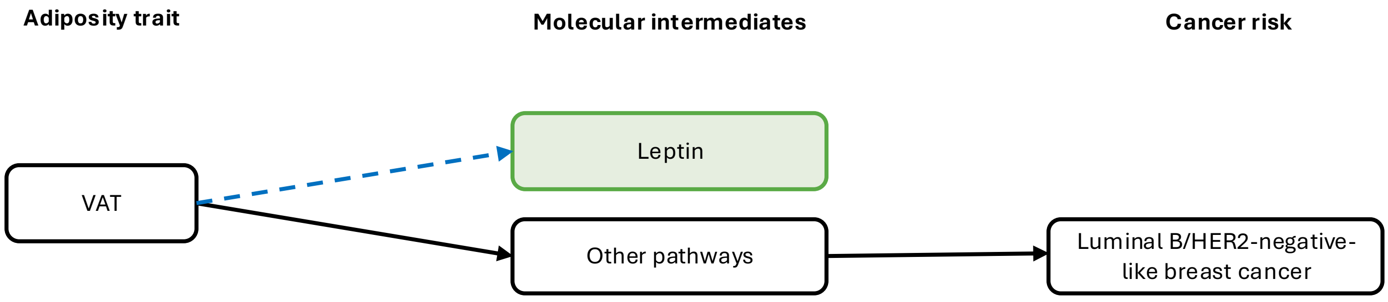

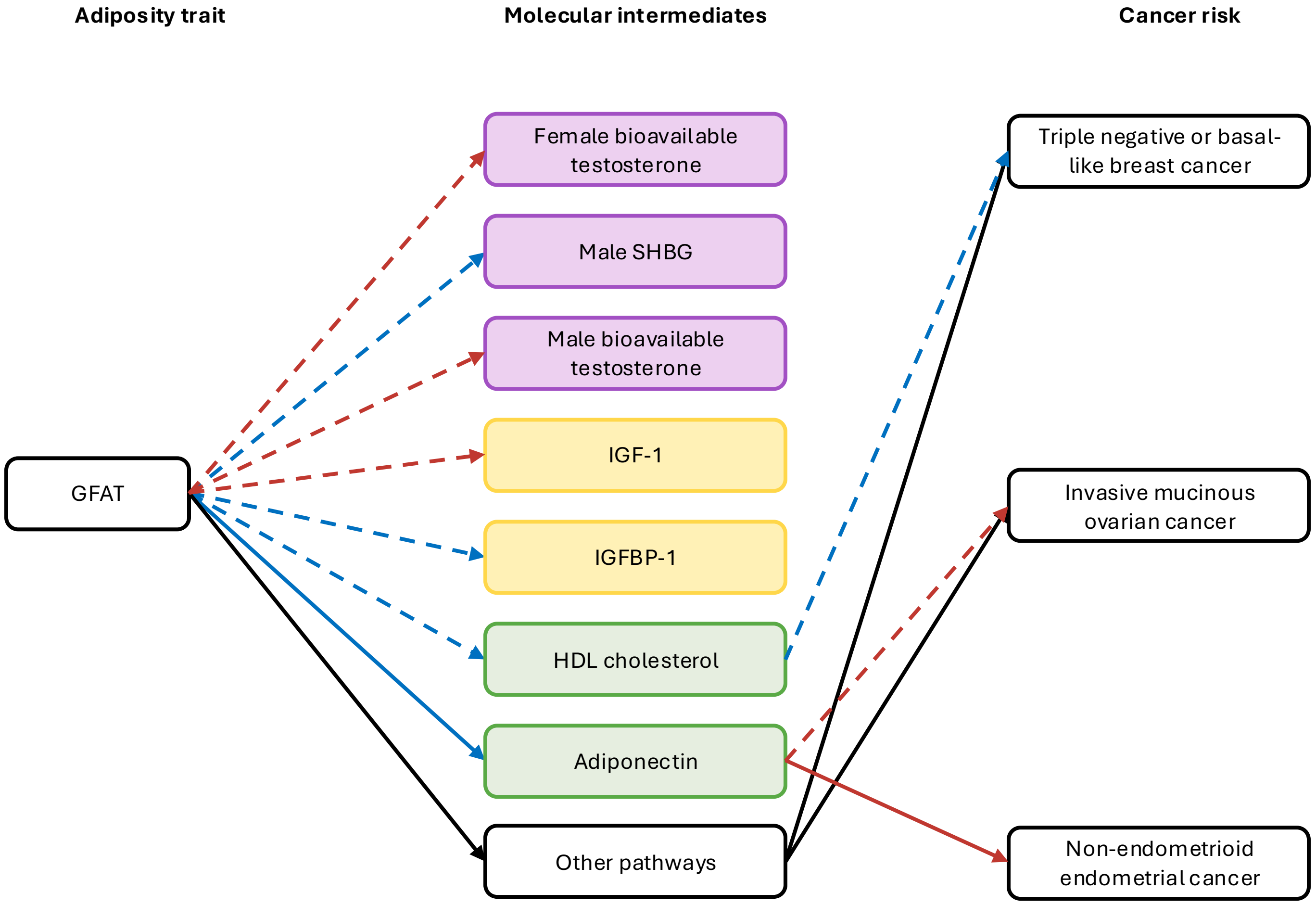

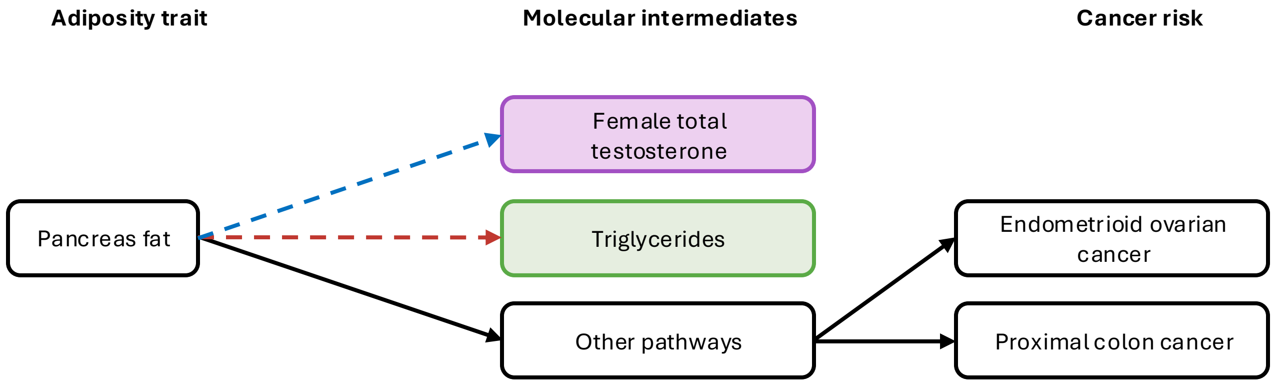

### Supplementary figure 1.png

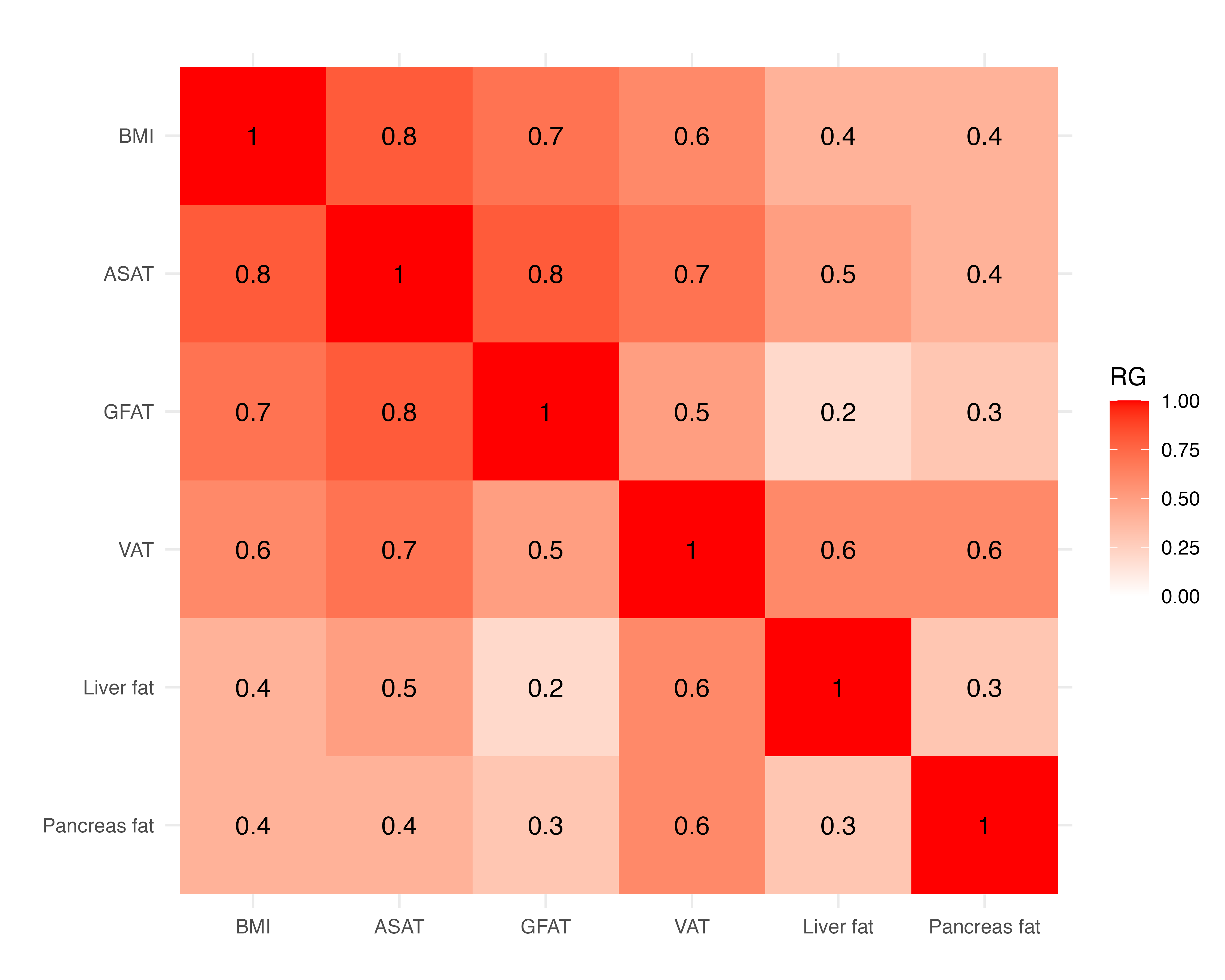

### Supplementary figure 2.png

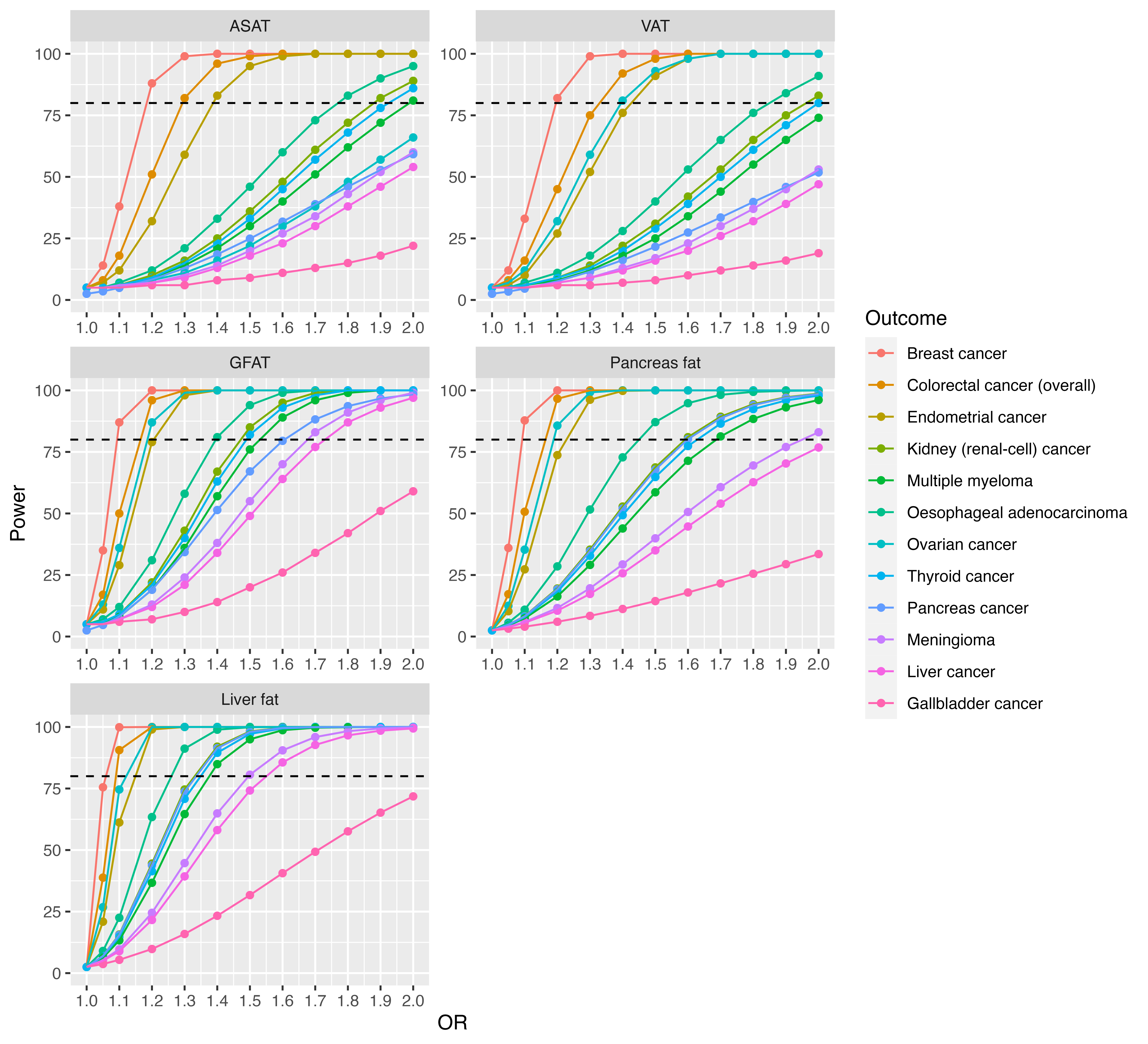

### Supplementary figure 3.png

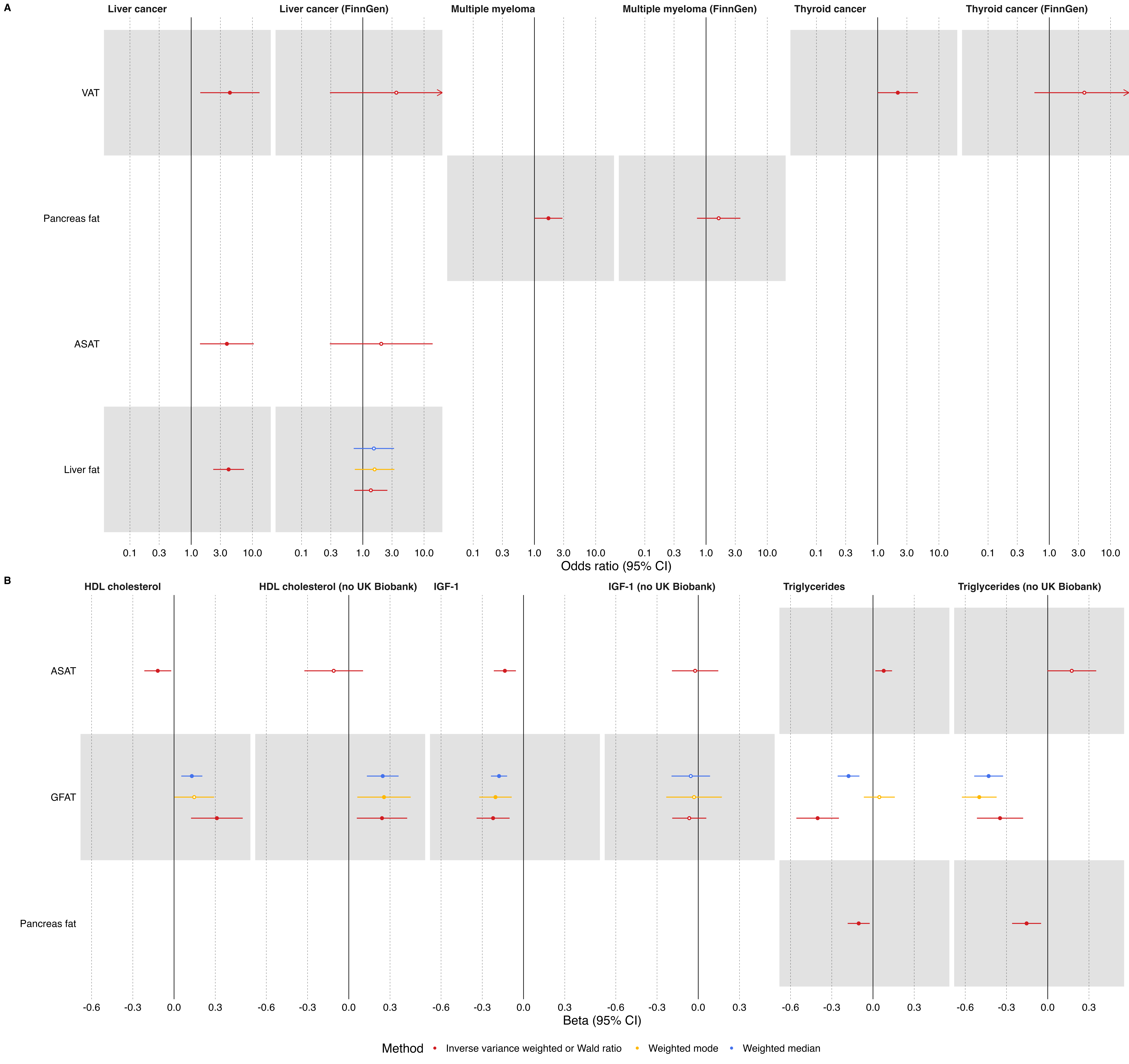

### Supplementary figure 4.png

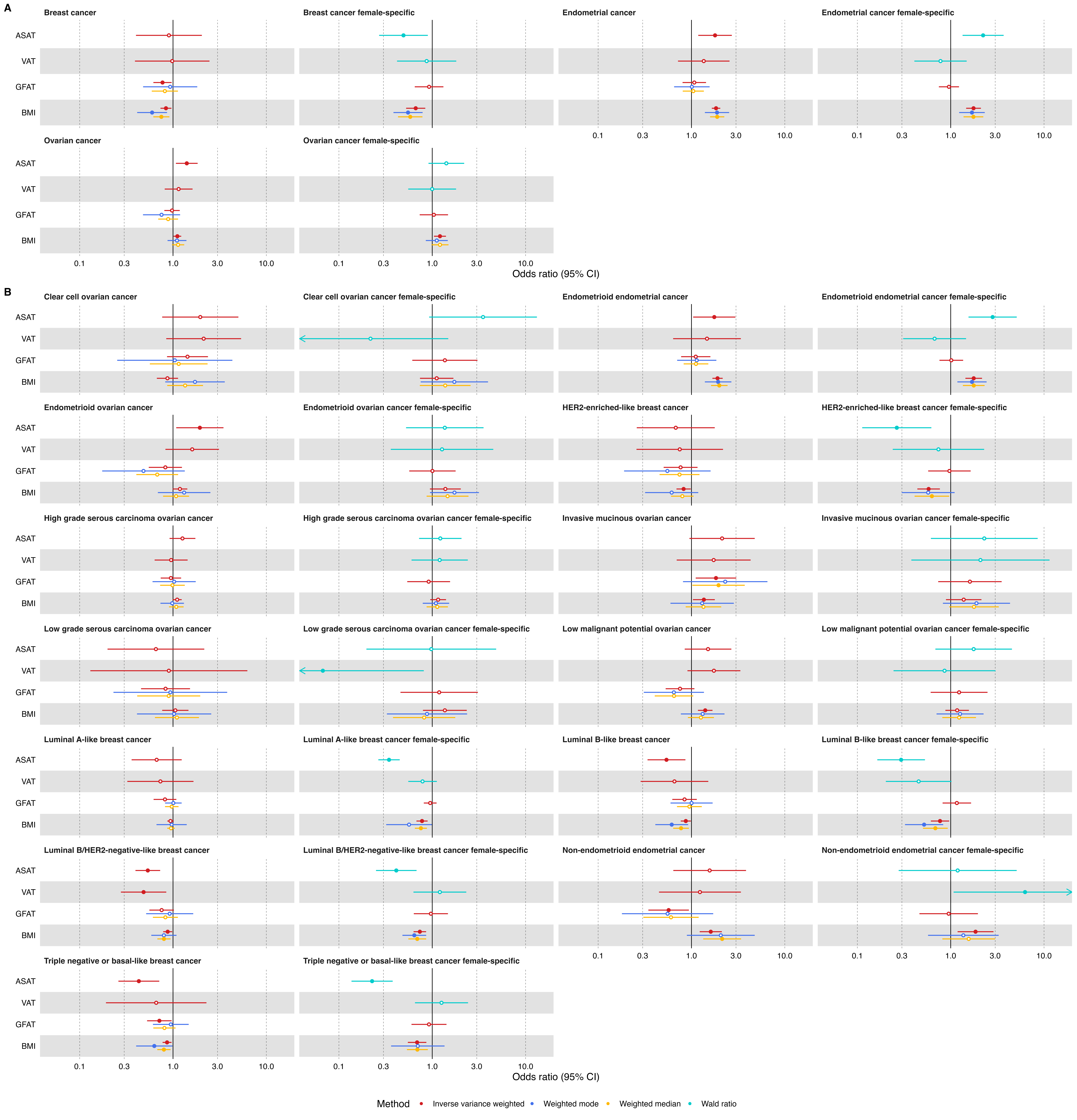

### Supplementary figure 6.png

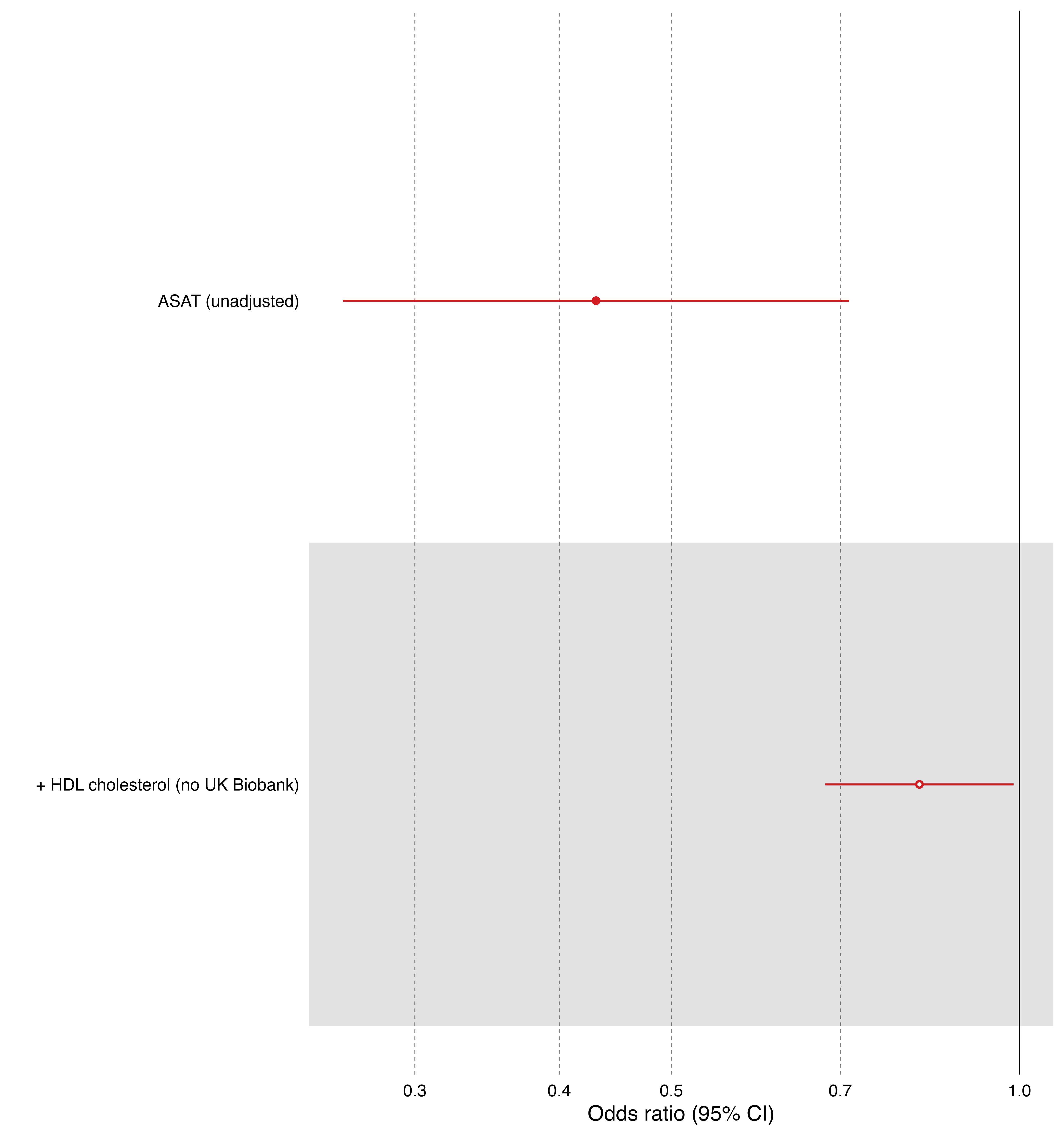

### Supplementary figure 7.png

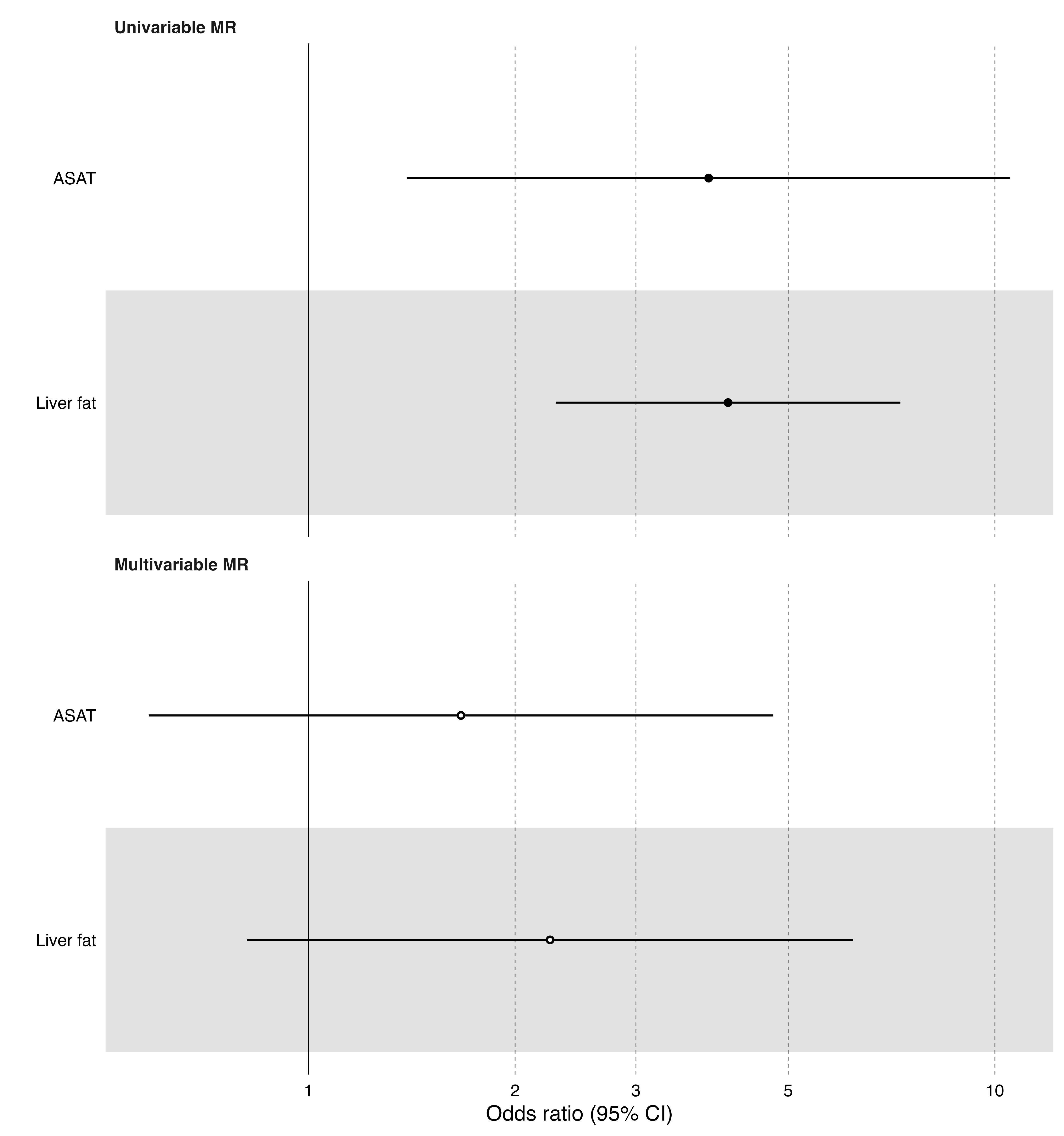
